# Predicting recovery outcomes following mild traumatic brain injury in an Australian community cohort: results from the Concussion Recovery Study (CREST)

**DOI:** 10.64898/2025.12.07.25341796

**Authors:** Jacinta Thorne, Jemma Keeves, Aleksandra K. Gozt, Gill Cowen, Elizabeth Thomas, Alexander Ring, HuiJun Chih, Caerwen Beaton, Francesca Buhagiar, Amanda Jefferson, Melissa Papini, Glenn Arendts, Antonio Celenza, Ben Smedley, Sjinene Van Schalkwyk, John Charles Iliff, Nicole Ghedina, Russell Young, Monica Marton, Suzanne Robinson, Melissa Licari, Dan Xu, Stephen Honeybul, Michael Bynevelt, Carmela Pestell, Daniel Fatovich, Sarah C. Hellewell, Melinda Fitzgerald

**Affiliations:** Curtin Medical Research Institute, Curtin University, Bentley WA, Australia; Perron Institute for Neurological and Translational Science, Nedlands WA, Australia; School of Allied Health, Faculty of Health Sciences, Curtin University, Bentley WA, Australia; School of Public Health and Preventive Medicine, Monash University, Melbourne VIC, Australia; Connectivity Traumatic Brain Injury Australia, Nedlands WA, Australia; Curtin Medical School, Faculty of Health Sciences, Curtin University, Bentley WA, Australia; School of Population Health, Curtin University, Bentley WA, Australia; School of Medicine, The University of Western Australia, Nedlands WA, Australia; Institute for Immunology and Infectious Diseases, Murdoch University, Australia; School of Psychological Science, The University of Western Australia, Nedlands WA, Australia; Discipline of Emergency Medicine, School of Medicine, The University of Western Australia, Australia; Emergency Department, Fiona Stanley Hospital, Murdoch WA, Australia; Emergency Department, Sir Charles Gairdner Hospital, Nedlands WA, Australia; Emergency Department, Rockingham General Hospital, Rockingham WA, Australia; Emergency Department, Joondalup Health Campus, Joondalup WA, Australia; Emergency Department, St John of God Hospital Murdoch, Murdoch WA, Australia; School of Medicine, The University of Notre Dame, Fremantle WA, Australia; Royal Flying Doctor Service - Western Operations, Australia; Emergency Department, St John of God Midland Public Hospital, Midland WA, Australia; Emergency Department, Albany Regional Hospital, Albany WA, Australia; Emergency Department, Geraldton Regional Hospital, Geraldton WA, Australia; Deakin Health Economics, Institute for Health Transformation, School of Health and Development, Deakin University, Melbourne VIC, Australia; The Kids Research Institute Australia, Nedlands WA, Australia; The First Affiliated Hospital, Sun Yat-Sen University, Guangzhou, China; Consultant Neurosurgeon, Sir Charles Gairdner Hospital, Royal Perth Hospital and Fiona Stanley Hospital, Perth WA, Australia; The Neurological Intervention & Imaging Service of Western Australia, Sir Charles Gairdner Hospital, Nedlands, Western Australia, Australia; Emergency Medicine, Royal Perth Hospital, Perth WA, Australia; Centre for Clinical Research in Emergency Medicine, Harry Perkins Institute of Medical Research, Nedlands WA, Australia

**Keywords:** mild traumatic brain injury, brain concussion, persisting post-concussion symptoms, recovery

## Abstract

**Background:** Most people recover well following mild traumatic brain injury (mTBI), however some experience persisting post-concussion symptoms (PPCS) for months or years. Our aims were (i) to evaluate the presence and impact of PPCS on return to functional activities over a 12-month period; and (ii) identify pre-, peri- and post-injury factors predictive of PPCS in an Australian community-based cohort at 3- and 12-months.

**Methods:** Adults (18–65 years) with mTBI were assessed by telephone within 7 days post-injury to capture demographics, pre-injury health status, injury circumstances (including mechanism, site of head impact) and symptoms. The primary outcome measure was the presence of PPCS, measured with the Post-Concussion Symptom Scale (PCSS). Logistic regression identified predictors of PPCS at 3- and 12-months.

**Results:** Of 232 participants, 49.7% had PPCS at 3-months and 45.6% at 12-months. Return to work was 96.7% at 3-months and 96.9% at 12-months, despite nearly 50% reporting PPCS. Participants with a high initial symptom burden (PCSS ≥30) were six-times more likely to experience PPCS at 3-months (adjusted odds ratio [aOR] 6.17, 95%CI 2.63–14.46) and three-times as likely to have PPCS at 12-months (aOR 2.70, 95%CI 1.01–7.23). Acute symptoms of “feeling slow” or “nervousness” predicted PPCS at 3-months (aOR 2.97 and 2.74, respectively), but not at 12-months.

**Conclusion:** Of those who completed follow-up, nearly half had persistent symptoms at 12-months, often continuing to work despite symptoms. High initial symptom severity and acute symptoms may help clinicians identify those needing early targeted follow-up to reduce the burden of PPCS.

## Introduction

Mild traumatic brain injury (mTBI), commonly referred to as concussion, is globally recognised as the most prevalent form of traumatic brain injury and a major public health issue(1). Mild traumatic brain injury can result from various mechanisms, including sports injuries, falls, transport-related incidents, assaults, and military-related blast events, and can affect individuals of all ages(2). Despite the term ‘mild’, many individuals experience persistent post-concussion symptoms (PPCS), involving physical, cognitive and psychosocial impairments that may last for months or even years after a mTBI, impacting their return to work (RTW), physical activity and study(3,4).

The timeframes and methodologies used to define PPCS in the literature varies widely, making it difficult to reach consensus on the true prevalence and standard recovery trajectory for individuals after mTBI(5). Using a Delphi consensus process, Lagacé-Legendre et al(6) defined PPCS as post-concussion symptoms that cannot be explained by pre-existing conditions and persist for three months or more after the injury, negatively impacting a person’s life – a definition that has since been adopted by multiple research groups(7–9). At three months post-mTBI, rates of PPCS have been reported to vary between 11 to 64%(8,10–12) with functional impairments continuing to impact nearly half of study cohorts at twelve months post-mTBI(4,13).

It is not well understood what proportion of people are able to RTW after mTBI but continue to experience PPCS. A delayed or unsuccessful return to functional activities such as work, physical activity, sport and study as a result of PPCS can have detrimental consequences on an individual’s financial, physical and psychosocial wellbeing(14,15). Despite more than 90% of people with mTBI reporting RTW within two months post-injury(14), nearly one fifth will experience productivity loss(16), a change in their employment status(17), reduced annual income(17) and limitations in their work capacity(16).

Evidence shows that early access to psychoeducation(11), targeted symptom-based interventions(18) and multidisciplinary community rehabilitation services can significantly improve recovery and reduce symptom burden following mTBI(19,20). Identifying the key predictors of poor outcome after mTBI is critical for developing risk stratification models and enabling early and targeted healthcare interventions for people identified to be most at risk of PPCS after mTBI to improve long term recovery outcomes and return to activities(3,21). Predictors identified in prior literature include a history of prior concussion(22), female sex(3,23,24), adolescence(23), mechanism of injury(25) and the presence of pre-existing conditions such as migraine(22,26), and mental health conditions(23,27). The severity and type of initial symptoms, particularly early post-injury dizziness, headache, and cognitive difficulties, are also strongly associated with a prolonged recovery(23,24,27). Psychological factors, including high levels of stress(28), poor coping mechanisms(28), and lower resilience(29) have been identified as contributors to persisting symptoms. Despite an improved understanding of the multi-factorial nature of recovery after mTBI, predictive models identifying people most at risk of a poor recovery have not been measured consistently across studies. Given the substantial short- and long-term impacts that persistent symptoms can have on an individual’s function and quality of life, there is a need to identify key predictors of outcome from broad community samples and so improve our understanding of recovery and inform follow up care.

The present study aims to (i) describe and evaluate the presence of PPCS, impact on quality of life (QoL) and ability to return to functional activities including work, physical activity, sport and study over a 12 month period following mTBI; (ii) compare return to functional activities (including work, physical activity, sport, and study) and QoL for people with and without PPCS at three and 12 months post-mTBI and (iii) to identify pre-, peri- and post-injury factors predictive of PPCS at three and twelve months post-injury in an all-cause community sample of people following mTBI.

## Methods

### Ethics Approval and Clinical Trial Registration for the Concussion REcovery STudy (CREST)

This study presents data collected for CREST - a prospective, longitudinal observational cohort study conducted across metropolitan and regional Western Australia between 5^th^ August 2019 and 31^st^ December 2023. Ethics approval for this study was obtained from Human Research Ethics Committees of Royal Perth Hospital (#RGS0000003024), Curtin University (HRE2019-0209), Ramsay Health Care (#2009) and St John of God Health Care (#1628). Recruitment commencement dates for each hospital site are as follows: Royal Perth Hospital 15^th^ August 2019; Sir Charles Gairdner Hospital 21^st^ August 2019; Fiona Stanley Hospital 4^th^ December 2019, Rockingham General Hospital 10^th^ December 2019, St John of God Hospital Midland 13^th^ February 2020, St John of God Hospital Murdoch 5^th^ March 2020, Joondalup Health Campus 6^th^ May 2020, Albany Regional Hospital 11^th^ December 2020, Geraldton Regional Hospital 11^th^ December 2020. All sites ceased recruitment 31^st^ December 2022. The Concussion REcovery STudy (CREST) was registered with the Australian New Zealand Clinical Trials Registry (ACTRN12619001226190).

### CREST Study Design

A detailed description of the CREST study protocol has been previously published(30), with a brief summary provided below. CREST recruited adults aged 18 to 65 years who had experienced mTBI within the previous seven days, from any cause of injury. Recruitment occurred via hospital Emergency Departments, as well as general practitioners (GP)/family doctors or self-referrals. All participants provided written consent for their contact details to be released to the study team following their medical review. Verbal consent was obtained by telephone prior to their participation in the study, and a copy of their verbal consent emailed to the participant. Upon enrolment, participants completed a semi-structured telephone interview, providing details on demographics, the injury event, acute mTBI management, previous history of mTBI, and relevant medical history. mTBI symptoms were assessed using the Post-Concussion Symptom Scale (PCSS), a 22-item inventory using a 7-point Likert scale for symptom prevalence and severity(31,32). The test–retest reliability of the PCSS measured as part of the ImPACT (Immediate Post-Concussion Assessment and Cognitive Testing) neuropsychological battery was 0.81(33).

Follow-up telephone interviews were conducted with participants at one, three, six, and twelve months after mTBI to document recovery experience and return to physical activity, sport, work, and study (if applicable). The PCSS was administered at each follow-up time point to assess ongoing mTBI symptoms. Where possible, follow-up telephone interviews were conducted by the same CREST research team member to maintain rapport with participants.

### Outcome measures

#### Post-Concussion Symptom Scale

The PCSS was used to evaluate symptoms and to assess recovery using a binarised cut-off score. An individual was deemed to be experiencing PPCS if they reported a PCSS symptom severity score of six or more for males, and seven or more for females(34) at three months or beyond following injury. This is consistent with other studies that have considered three months as a clinically significant time point for diagnosing PPCS(6–8).

#### Quality of Life

QoL was evaluated using the short form, six-item Quality of Life after Brain Injury – Overall Scale (QOLIBRI-OS)(35). The QOLIBRI-OS consists of six domains of health-related quality of life relevant following brain injury – namely, physical condition, cognitive function, emotions, autonomy with daily activities, social relationships and future prospects(35). It has good test-retest reliability (0.81) and high correlation with the full QOLIBRI scale (r=0.87)(35). Responses were converted to a percentage score and dichotomised using a recommended cut score of 52% to indicate a better (≥52%) or impaired quality of life (<52%)(36–39).

#### Return to physical activity, sport, work and study

At each follow-up interview, participants were asked if they had returned to physical activity, sport, work and/or study (if applicable), and if so, how many days following mTBI they returned to each activity. Further information regarding the type of physical activity or sport undertaken, and type of work were also collected.

### Statistical Analysis

Data analyses were conducted using Stata Statistical Software, Version 18 (Stata Corp LLC, Texas, USA), and figures were generated using GraphPad Prism 9.0 for Windows (GraphPad software, San Diego, California). Statistical significance was set at p<0.05 for all analyses.

For the first aim of the study, descriptive statistics were used to summarise participant demographic, pre-injury and injury-related characteristics for the cohort. Median and interquartile ranges (IQR) were provided for continuous data and counts and percentages for categorical variables. Outcome measures including the presence of PPCS, impact on QoL, and return to functional activities including work, physical activity, sport and study were reported for each timepoint (one, three, six and twelve months) to provide a descriptive overview of recovery trajectories.

For the second and third aims, all analyses focus on the three and twelve month timepoints. The relationship between PPCS and each functional outcome (return to work, physical activity, sport and study), and PPCS and QoL, was assessed using chi-squared tests or Fisher’s exact test when expected cell counts were less than five. Mann Whitney U tests were used to assess differences in QOLIBRI-OS total scores between those who met the criteria for PPCS and those who had recovered at each timepoint.

Addressing the third objective of this study, participant sociodemographic, pre-injury, peri-injury and clinical characteristics were assessed with respect to the main outcome of interest (PPCS or no PPCS) using exploratory univariate logistic regression analyses at three and twelve months. Further elaboration of included variables is provided as follows: socioeconomic status was determined using the Index of Relative Socioeconomic Disadvantage from the Australian Bureau of Statistics(40), a general socio-economic index based upon participant residential postcode; past history of mTBI was reported using categories of none, 1-2, and 3 or more; pre-injury diagnoses of mental health issues (self-reported in the demographic survey) including anxiety, depression and other psychiatric disorders were combined into one variable; regular prescription medication use was included to provide a general indicator of co-morbid health conditions; regular exercise (more than 3 times per week) as an indication of general fitness(41); mechanism of injury was reported as sport or non-sport related; concurrent fracture was used as an indication of severity of co-occurring injuries, with some evidence that fractures may also interact with brain injury(42). The association between location of head impact and PPCS was examined in two ways: firstly, by examining each individual site of head impact separately; and then comparing those who had one point of impact to those who had multiple or no reported impact to the head.

Symptom presentation results from the PCSS were represented in the following ways:

i. total symptom scores at enrolment - number of symptoms (out of 22) and symptom severity score (out of 132).
ii. binarised using the median as the cut-off point to enable ease of interpretation and clinical utility as a predictor of outcome in a clinical setting (PCSS number of symptoms - 0 to 11, 12 or more; PCSS symptom severity score 0-29, 30 or more).
iii. symptom clusters (physical - headache, nausea, vomiting, balance, dizziness, visual problems, fatigue, light sensitivity, noise sensitivity, numbness, cognitive - foggy, feeling slow, difficulty concentrating, difficulty remembering, emotional - irritable, sadness, more emotional, nervousness, sleep - drowsiness, sleeping less, sleeping more, trouble falling asleep)(43). Scores for each symptom cluster were developed by summing each symptom endorsed (yes=1/no=0) in that group at initial enrolment.
iv. individual symptoms were analysed separately with each symptom as a binary variable (yes/no). This variable was included based upon feedback that clinicians tend to only ask about whether a symptom is present or absent, and do not often use validated scales.

Variables identified as promising predictors of PPCS (p<0.05 on univariate analysis) were then included in multivariable modelling using the ‘enter method’(44) to allow for inclusion of all candidate predictors in the model simultaneously. Variables with only a small number of responses (less than 10% of total responses) were not included in ongoing analyses (pre-injury sleep disorders, ADHD, alcohol consumption at time of injury). Symptom presentation was used as the primary independent variable of interest and tested in separate multivariable models using the structures outlined previously.

Model performance was first evaluated using pseudo-R^2^ values, and then discrimination assessed using receiver operating characteristics (ROC) analyses to calculate area under the curve (AUC). Sensitivity, specificity, and correct classification values were also calculated to evaluate model performance.

#### Assessment of Bias

The potential for selection bias was assessed by comparing demographic characteristics (age and sex) of eligible participants who were referred to the study but not subsequently enrolled to those who were enrolled using descriptive statistics. Attrition bias was assessed by comparing those participants who completed all follow-up timepoints (1, 3, 6 and 12 months) to those who had at least one follow-up missing, across a range of demographic and clinical variables(45). Any statistically different factors identified from this analysis were included as covariates in subsequent multivariable modelling to account for their potential effect.

## Results

### Description of the cohort

Participant demographic information, injury-related characteristics and prior health history for the cohort (n=232) are summarised in Table 1 with additional data provided in Supplementary Table 1. Participants had a median age of 33 years, 43.5% were female, and were enrolled a median of four days following mTBI (Table 1). Participants were primarily referred to CREST via hospital emergency departments across metropolitan (Perth) and regional Western Australia (94.8%, n=220), with the remainder referred from general practitioners or self-referred following medical diagnosis (5.2%, n=12). Approximately one third of participants (31.9%) sustained their mTBI during sport, with the remainder from non-sport related activities. 28.4% (n=66) of participants had experienced loss of consciousness (LOC) associated with their mTBI, and 50.4% (n=116) experienced post-traumatic amnesia.

**Table 1:**
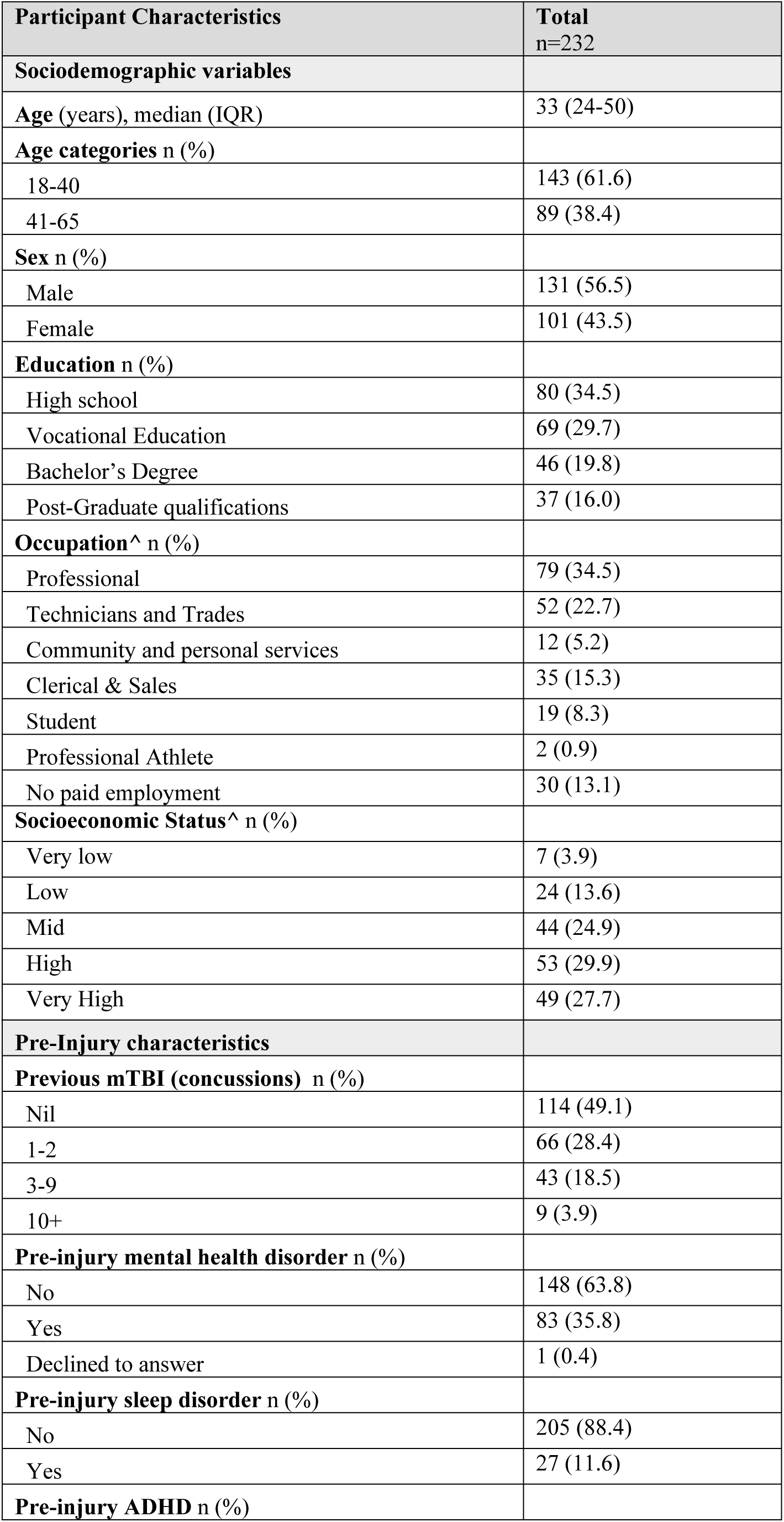
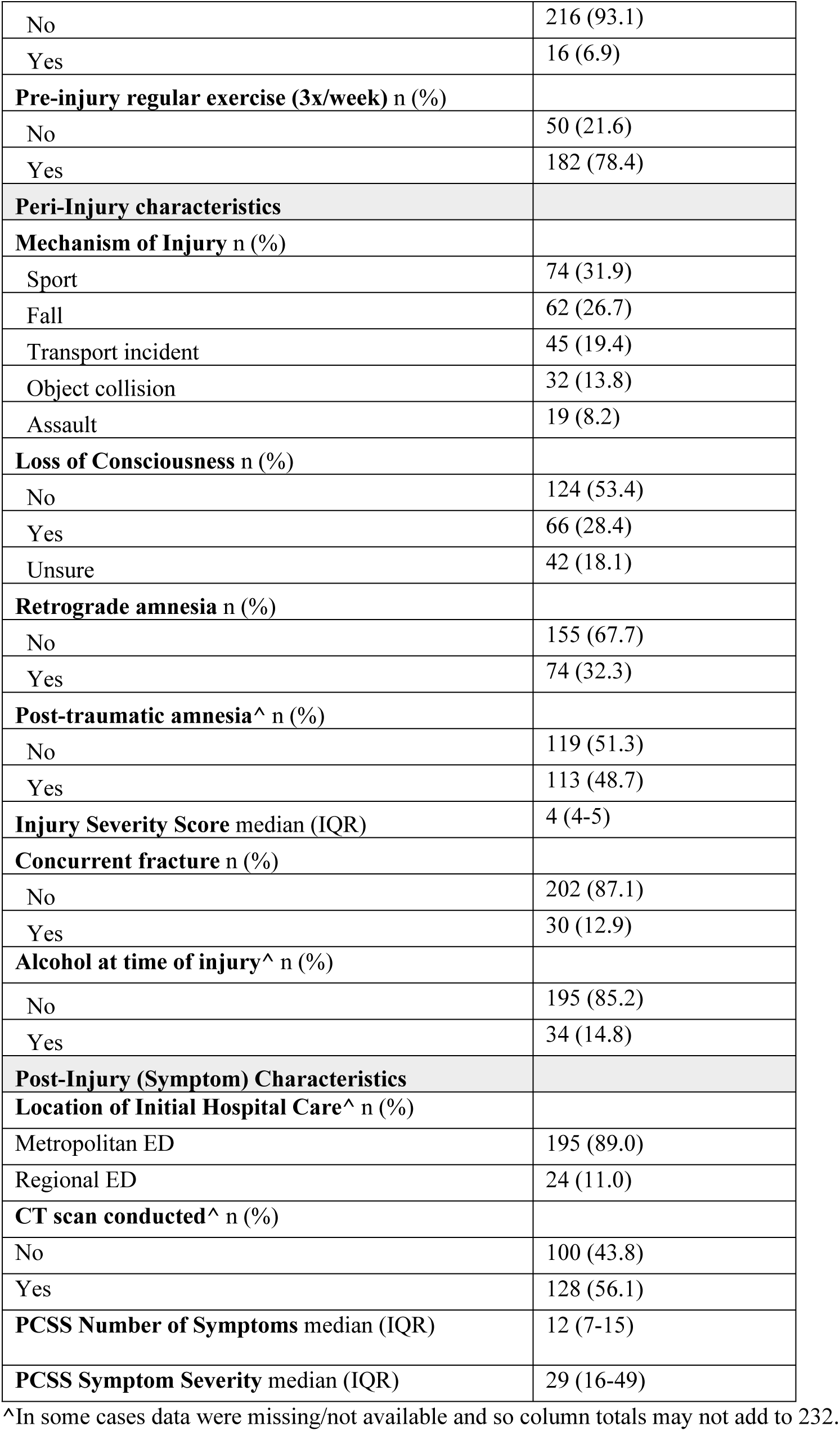
Demographic, pre-injury and injury-related characteristics for the CREST cohort (n=232)

A further 208 participants were referred to the study but were not subsequently enrolled. 72 of these participants did not meet the inclusion criteria, and the remaining 136 participants who were eligible could not be contacted or declined to participate. The demographic characteristics of these participants are provided in Supplementary Table 2, but were comparable to the main cohort (median age 29 years, 39% female).

### Recovery outcomes over time

Follow up data were acquired for 186 (80.2%) of participants at one month, 165 (71.1%) at three months, 150 (64.6%) at six months and 138 (59.5%) at twelve months post mTBI. Given the attrition rate to follow up, further analyses were conducted to compare those who completed all four follow-ups timepoints (n=94, 40.5%) to those who missed at least one follow-up. No significant differences in demographic or injury-related factors, or severity of initial injury were identified between the groups (Supplementary Table 2). However, the group who completed all four follow-ups were found to have a significantly lower level of pre-existing mental health issues (p=0.03).

#### Symptom burden

At enrolment in the study (4.3±1.7 days following injury), the median number of symptoms reported was 12 (IQR 7-15), with a median symptom severity score of 29 (IQR 16-49, range 0-108)). By three months, symptom severity scores had decreased to a median of 6 (IQR 0-19) and remained relatively similar until twelve months (median = 5, IQR 0-16). Median symptom severity was higher for females than males at all timepoints, with a significant difference seen initially (p<0.001), and at one month (p=0.01) and three months post mTBI (p=0.004) (Figure 1a). No differences were noted between females and males at six or twelve months. Compared to sport-related mTBI, people who sustained a mTBI from a non-sport related incident reported significantly higher symptom severity scores initially (p=0.004), and at one and three months (p=0.003 and p<0.001 respectively), although symptom severity scores were comparable at six and twelve months (Figure 1b).

**Figure 1.**
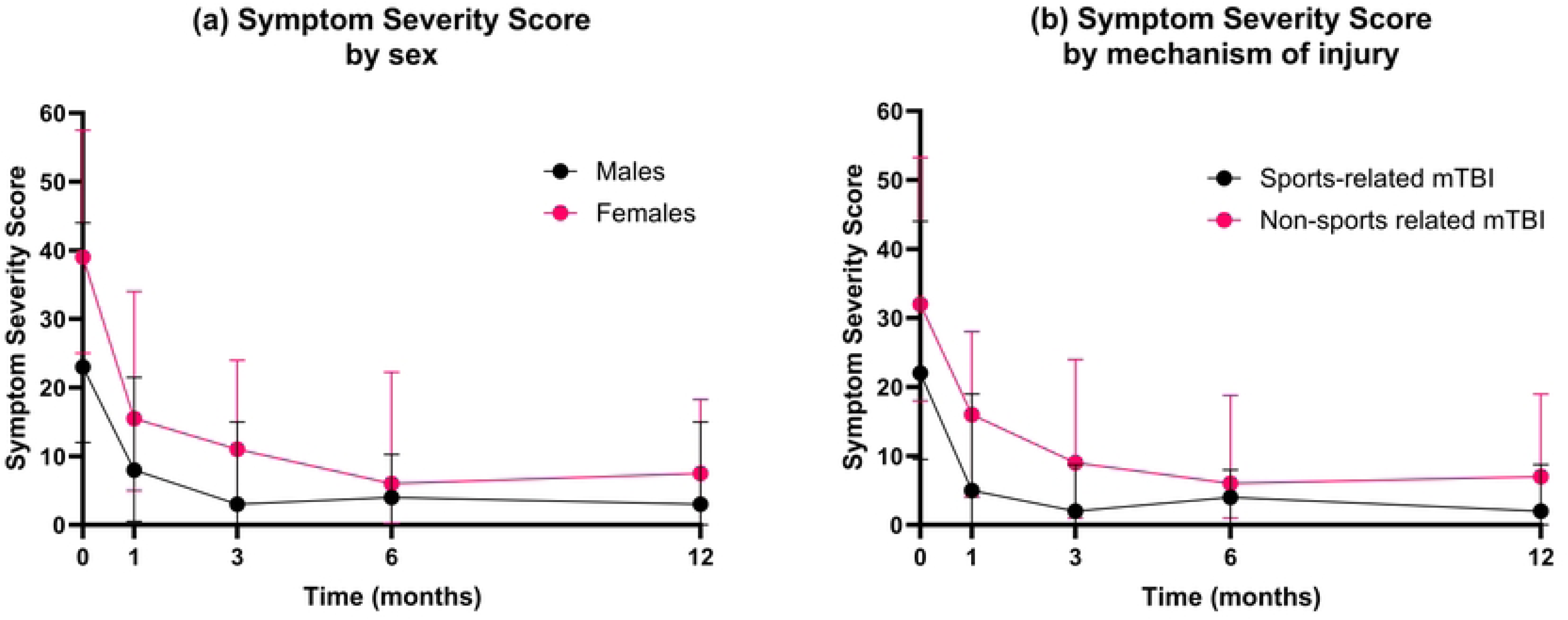
PCSS Symptom Severity Score over time for all participants, according to (a) sex and (b) mechanism of injury (sport vs non-sport related mTBI)

#### PPCS over time

At one month post-injury 114 (61.3%) participants were still experiencing persisting symptoms. By three months this reduced to 82 (49.7%), and then remained relatively consistent at six (n=69, 45.7%) and twelve months (n=63, 45.6%). The presence of PPCS at three and twelve months was also explored for individual participants using a Sankey diagram (Figure 2). At twelve months, most participants who reported PPCS at 3 months remained symptomatic, though some recovered. Conversely, while many participants without PPCS at 3 months stayed symptom-free, a notable group (n=23) reported PPCS when assessed at twelve months. These patterns highlight both the persistent and variable nature of PPCS after mTBI.

**Figure 2.**
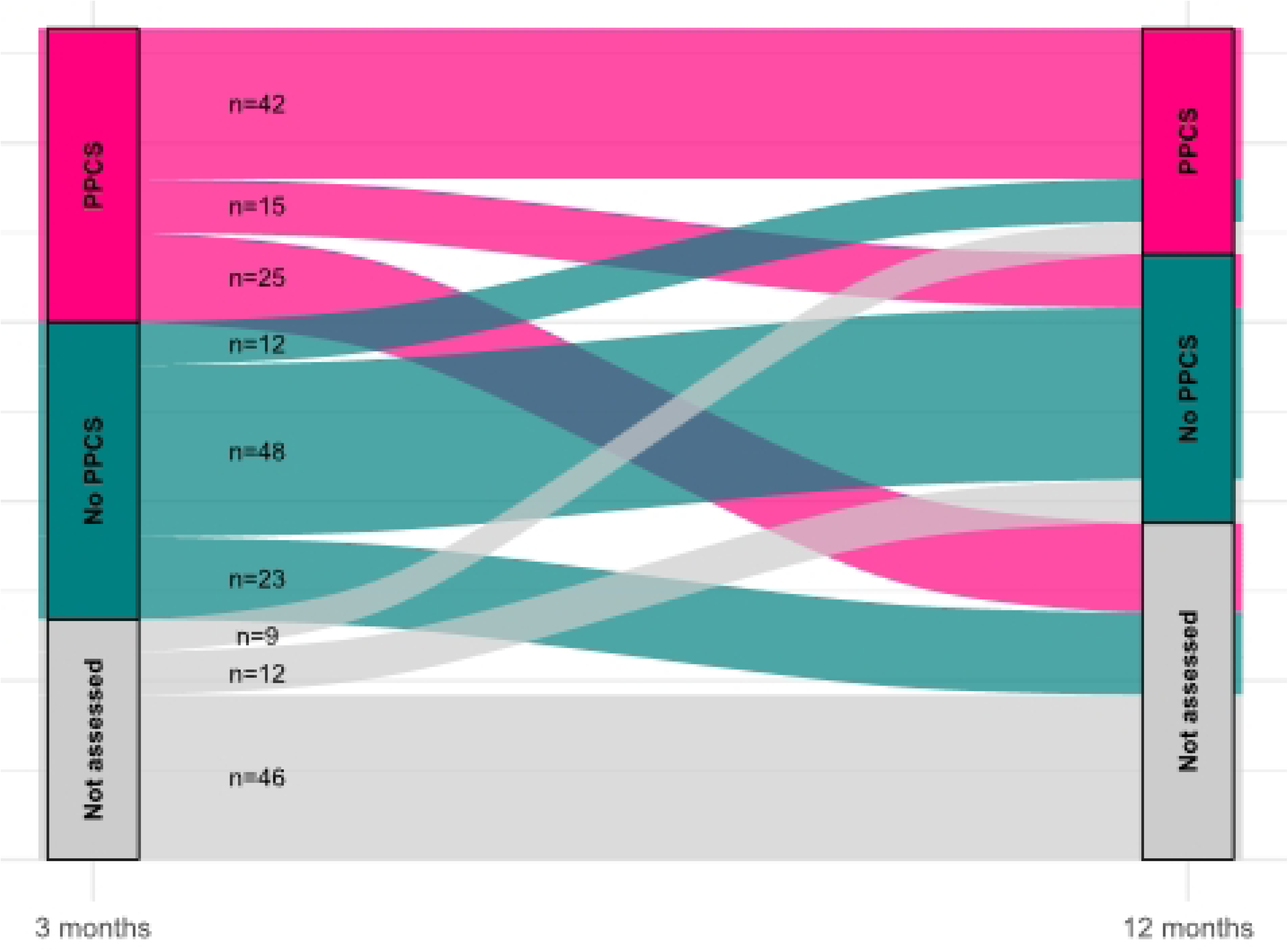
Change in PPCS status between three and twelve months post-injury. Each flow represents the number of participants moving between categories: PPCS (pink), No PPCS (teal), and Not assessed (grey). Counts (n) are shown alongside each flow

#### Individual symptoms over time

At the time of enrolment, fatigue was the most frequently reported symptom (78.9%, n=183), followed by headache (n=170), and then cognitive symptoms including feeling slowed down (n=169), difficulty concentrating (n=165), and feeling foggy (n=162). The least frequently reported symptom was vomiting (n=7).

Individual symptom presentation over time for participants who completed all four follow-ups (n=94) is displayed in Figure 3. The cumulative frequency for each symptom provides an indication of symptom burden over time, with fatigue, difficulty concentrating and remembering, and irritability remaining pronounced over the twelve month period. In contrast, some symptoms (i.e. dizziness, balance problems and nausea) appear to become less prominent with time.

**Figure 3.**
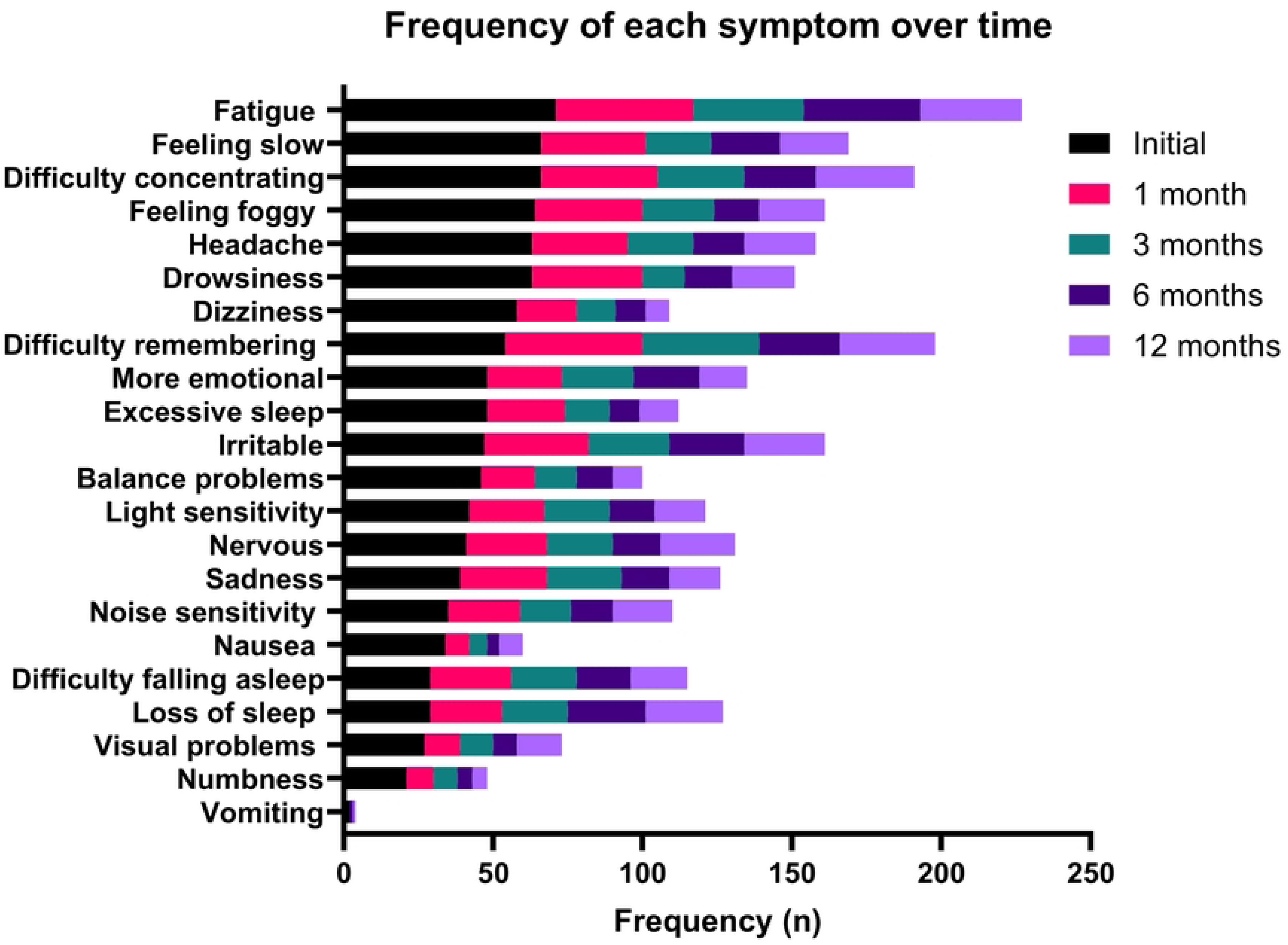
Cumulative frequency of individual symptoms at each timepoint shown in order of most to least frequently reported at initial enrolment. Data shown is for participants who completed all follow-up timepoints (n=94)

#### Return to functional activities

The proportion of participants who had returned to various functional activities at each follow-up timepoint is illustrated in Figure 4. As not all people participated in each functional activity pre-injury, results are expressed as a percentage of those who returned to their pre-injury baseline activities. Of those working pre-injury, the median for RTW was 7 days (IQR 3-11). Most participants (90.5%) had returned to work by one month. By three months, 96.7% had returned to work and this remained consistent at six and twelve months (90.4% and 97% respectively), Table 2.

**Figure 4.**
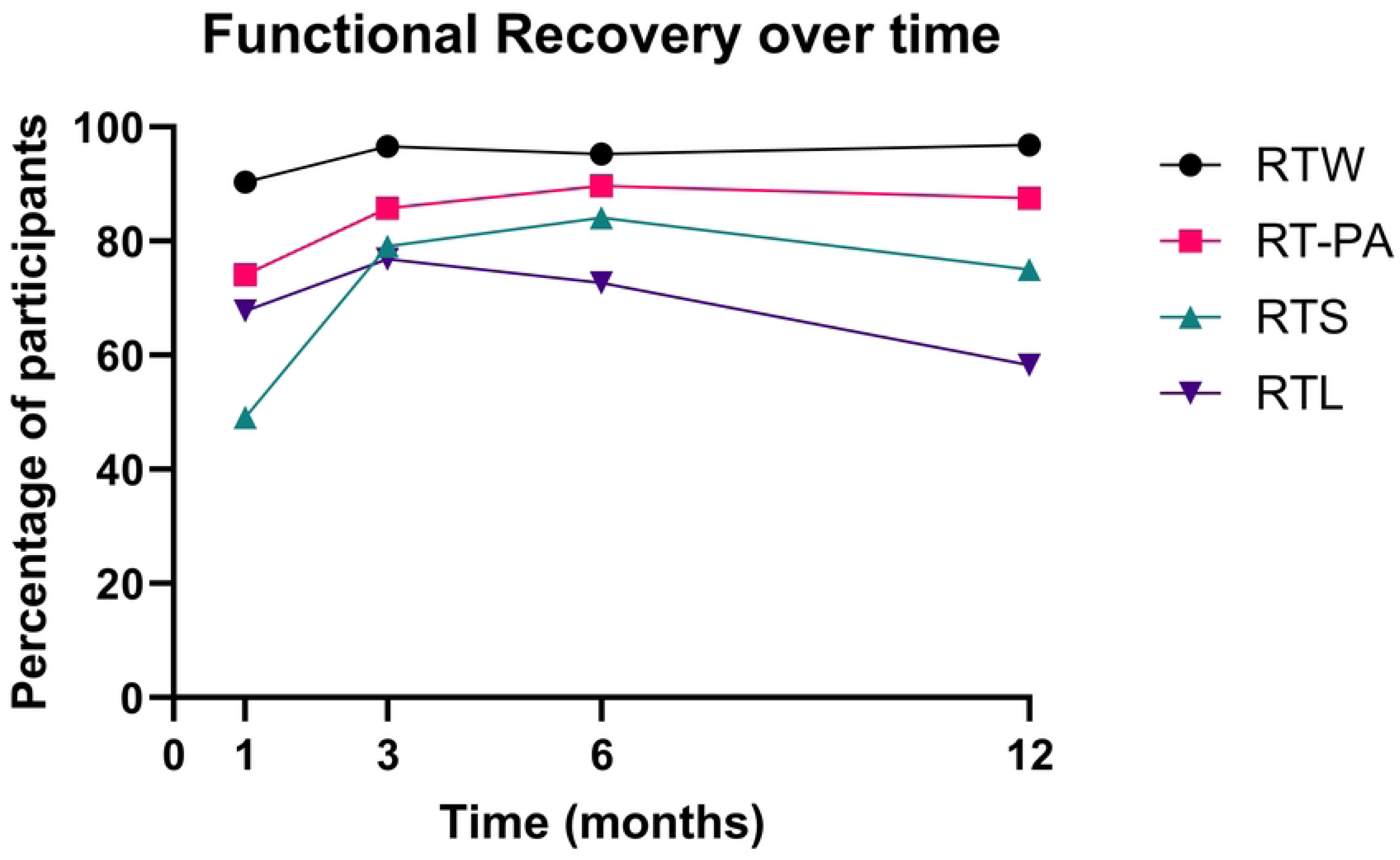
Percentage of participants who had returned to each functional activity at each timepoint (one, three, six and twelve months) post-injury. Abbreviations: RTW – return to work, RT-PA – return to physical activity, RTS – return to sport, RTL – return to learn (study). Percentage values were calculated as a proportion of those who had returned to an activity from their pre-injury status, and thus the total number of participants for each activity varies

**Table 2:**
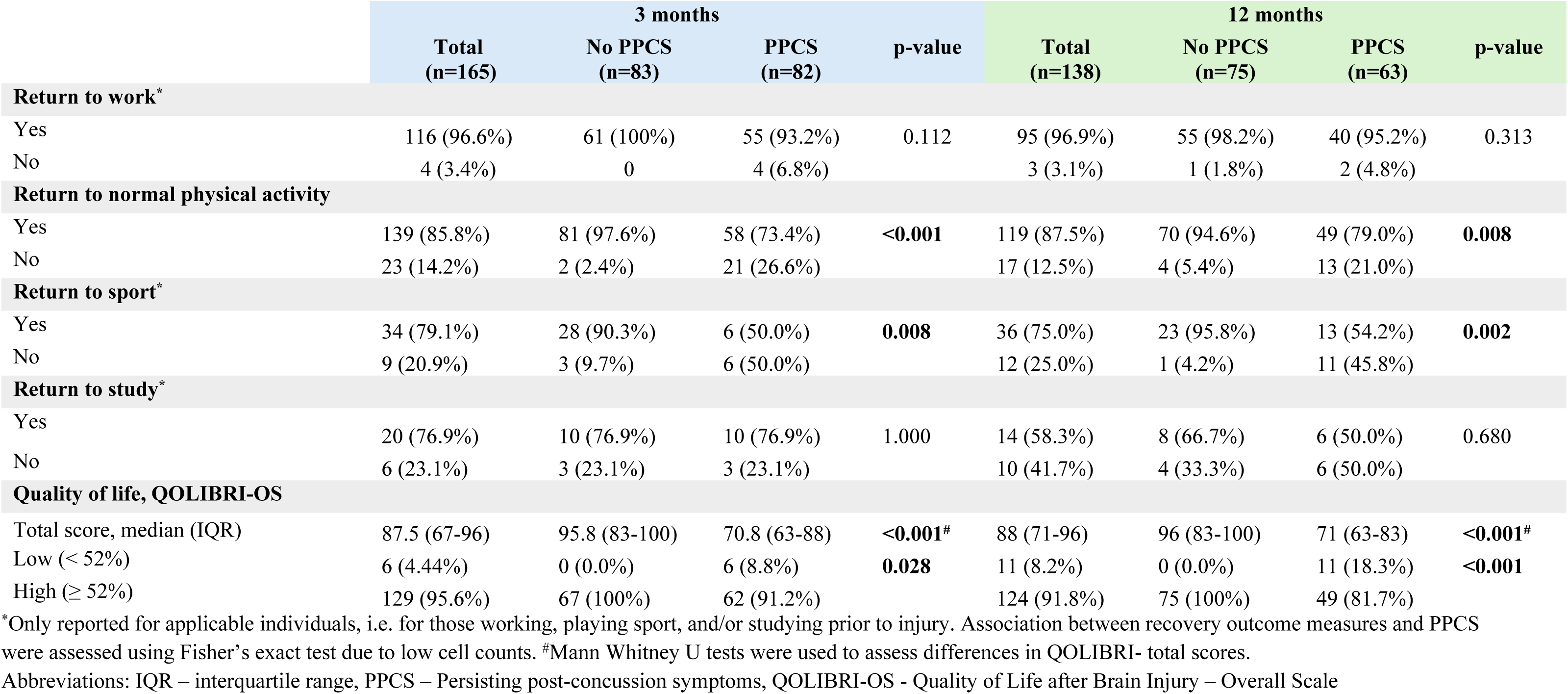
Summary of recovery outcome measures and the presence of PPCS at 3 and 12 months following mTBI.

Similar trends were observed for return to physical activity, with 74.1% returning to some form of exercise by one month, increasing to 85.8% by three months and remaining similar at six and twelve months. In contrast, only 49.1% had returned to sport at one month. This increased to 79.1% of participants at three months, 84.1% at six months and 75.0% at twelve months.

Of those participants who were studying prior to injury, 67.8% had returned to study by one month post-injury. Whilst there was an initial increase to 76.9% of participants at three months and 72.7% at six months, a decrease was then seen in the proportion of participants studying at twelve months (58.3%). Data were not available to determine whether these differences were attributable to mTBI or to other factors such as change in study status (e.g. semester enrolments, course completion).

### Relationship between functional recovery, QoL and PPCS outcomes

Nearly half of all participants from this all-cause mTBI sample continued to experience PPCS at both three months (49.7%) and twelve months (45.7%) post-injury. Of those working pre-injury, 96.6% had RTW by three months post-injury, but notably, 47.8% were still experiencing PPCS (Table 2). At twelve months, 96.9% of people working pre-injury had RTW, with 42.1% working with PPCS (Table 2). At three months, all participants who had not returned to work were employed in ‘Technician or Trade’ roles (n=4). The presence of PPCS was not associated with RTW at three or twelve months post-mTBI, Table 2.

At three months and twelve months post mTBI, significantly more participants with PPCS (26.6% and 21.0%) had not returned to normal physical activity compared to those without PPCS (2.4% and 5.4%) (Table 2). At three months, 40% of respondents (n=20) reported that they were less physically active as a result of their mTBI; however, this reduced to 25% at twelve months.

Of participants who played sport prior to their injury, 79.1% had returned to sport by three months post mTBI, with only 17.6% experiencing PPCS (Table 2). Significantly fewer people returned to sport at three months if they had PPCS compared to those without PPCS (50.0% v 90.3%, p=0.008). A similar association between return to sport and PPCS was also seen at twelve months post-mTBI (p=0.002) (Table 2).

Twenty-six participants reported studying at the time of injury. No significant association was found between returning to study and experiencing PPCS at either three months or twelve months post mTBI (p = 1.000 and p=0.680 respectively).

People with PPCS reported significantly lower QoL scores on the QOLIBRI-OS at both 3-months and twelve months post mTBI (p < 0.001) compared to those not experiencing PPCS. When considered as a binary outcome (low or high QOLIBRI-OS scores), a significantly higher percentage of people with PPCS had a low score at three months (17.6% vs. 0%, p < 0.001) and twelve months (23.3% vs. 0%, p < 0.001) post mTBI compared to those without PPCS (Table 2).

### Relationship between predictive factors of interest and PPCS

#### Demographic factors

Initial univariate exploratory analysis of potential predictors of PPCS at three and twelve months post mTBI is provided in Table 3. Older age (OR 1.98, 95%CI 1.05-3.71, p=0.033) and female sex (OR 2.74, 95%CI 1.43-5.23, p=0.002) were both significant predictors of PPCS at three months. Participants with a vocational level of education were four times more likely to experience PPCS relative to those with high school level education (OR 4.29, 95% CI 1.88-9.78, pp=0.001) at three months. However, those with university level education were not significantly more likely to experience PPCS. The same associations between demographic variables and PPCS were not apparent at twelve months. No significant associations were identified between socioeconomic status and PPCS at either timepoint.

**Table 3:**
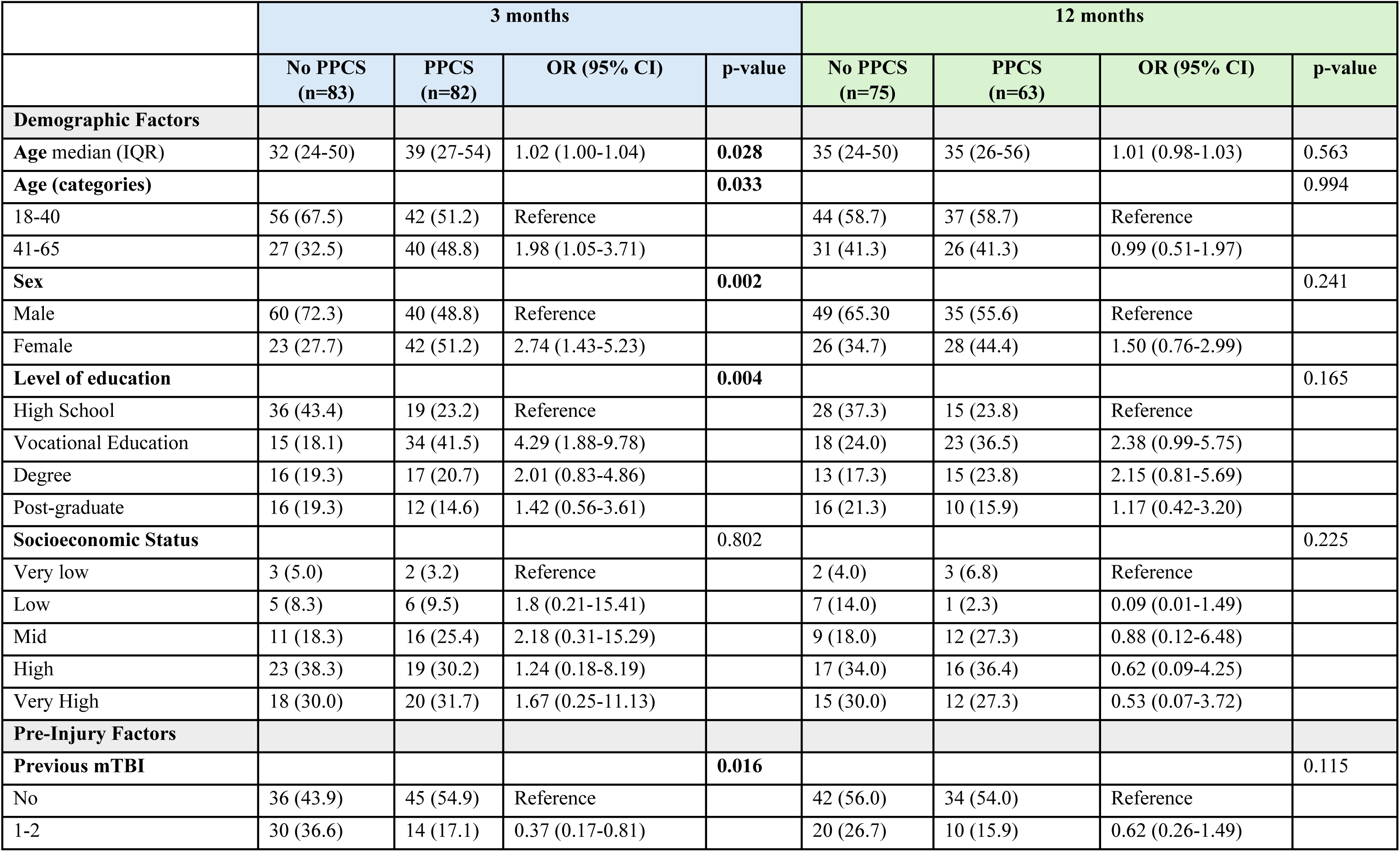
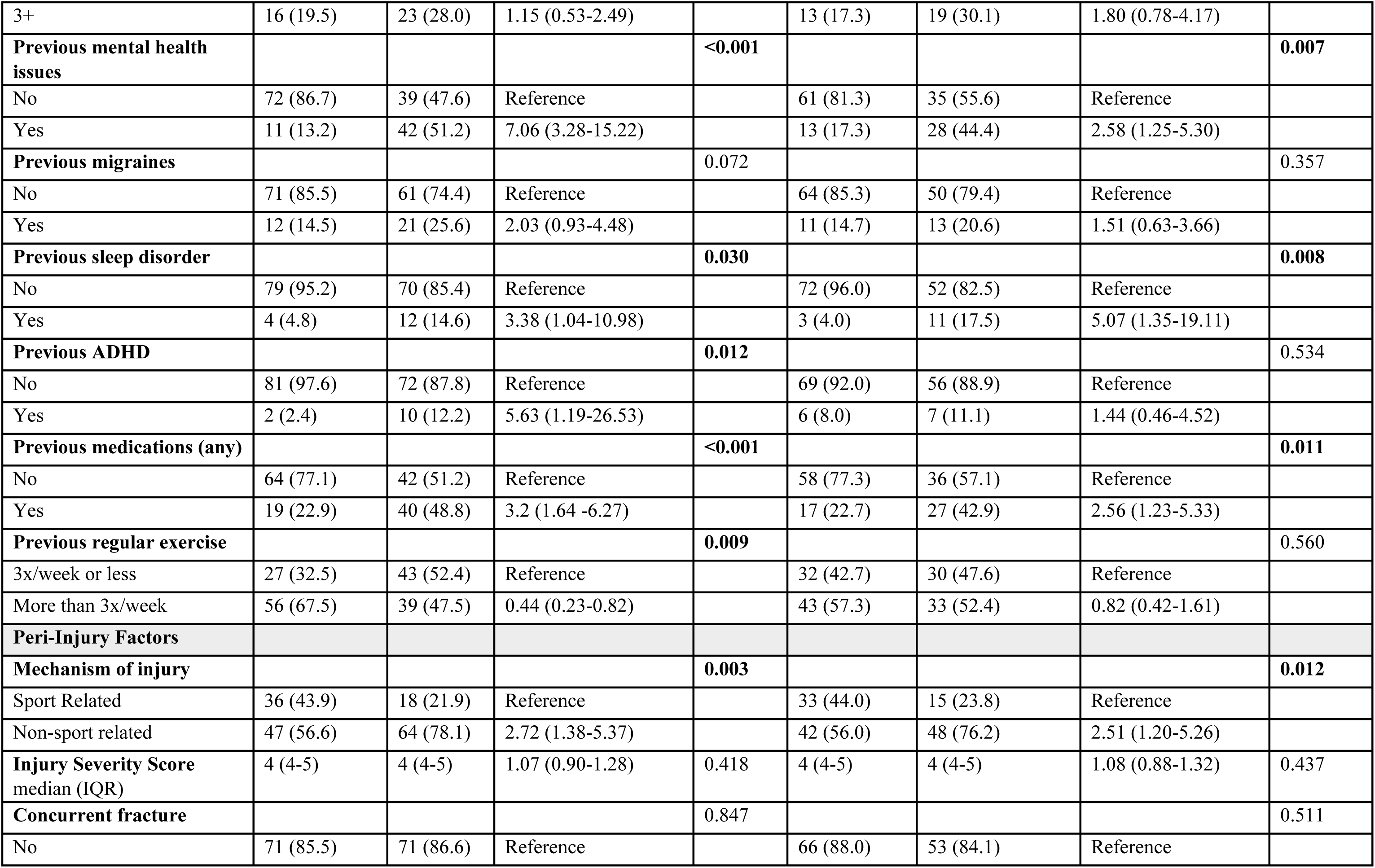
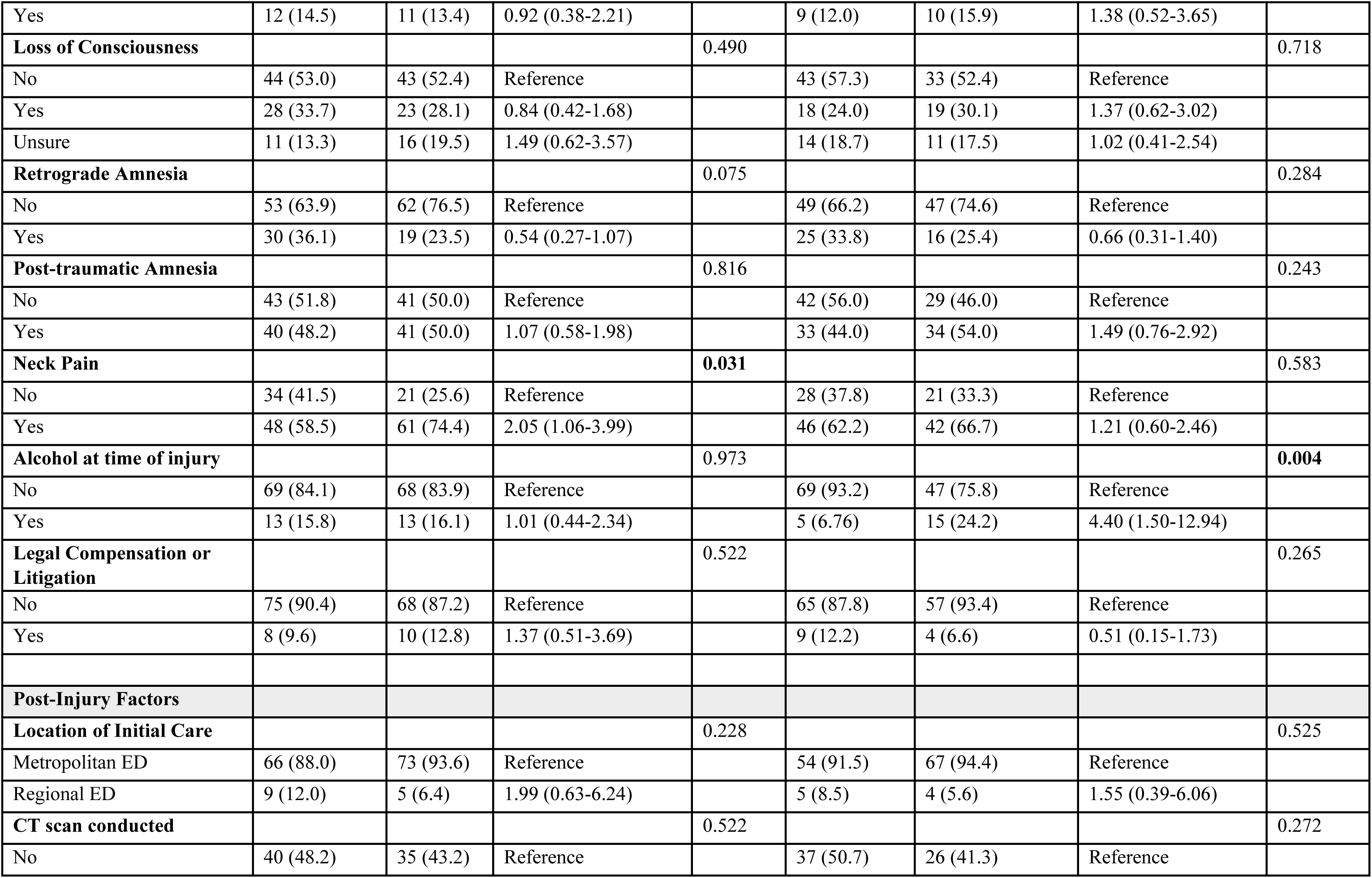
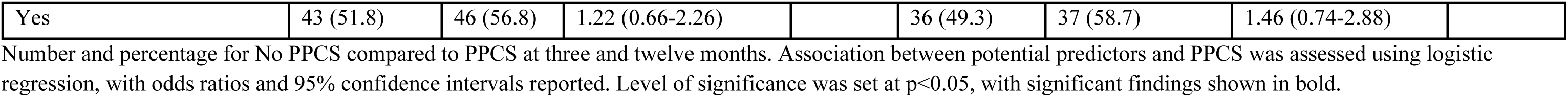
Exploratory univariate analyses of potential predictors of PPCS at 3 and 12 months following mTBI.

#### Pre-injury factors

Several pre-injury factors were associated with PPCS at three and twelve months (Table 3). Participants who reported a pre-injury diagnosis of mental health issues (depression, anxiety or other psychiatric disorders) had seven times the odds of PPCS at three months (OR 7.06, 95%CI 3.28-15.22, p<0.001), and were 2.5 times as likely to experience PPCS at twelve months (OR 2.58, 95%CI 1.25-5.30, p=0.007). Pre-existing sleep disorder was also associated with PPCS at both three and twelve months (OR 3.38, 95% CI 1.04-10.98, p=0.03; OR 5.07, 95% CI 1.35-19.11, p=0.008 respectively). Of interest, those who had experienced 1-2 prior mTBI’s were *less* likely to experience PPCS at three months compared to individuals with no history of previous mTBI (OR 0.37, 95% CI 0.17-0.81, p=0.016), whilst those who reported three or more previous mTBI were more likely to have PPCS (OR 1.15, 95% CI 0.53-2.49, p=0.016). Those who reported engaging in regular exercise pre-injury were less likely to experience PPCS at three months (OR 0.44, 95%CI 0.44, p=0.009) than those who did not exercise regularly. This association was not apparent at twelve months.

#### Peri-injury factors

In contrast, fewer peri-injury factors were associated with PPCS at three and twelve months (Table 3). Those who had sustained mTBI from non-sport related incidents were more than twice as likely to experience PPCS at both three (OR 2.72, 95% CI 1.38-5.37, p=0.003) and twelve months (OR 2.51, 95% CI 1.20-5.26, p=0.012). The presence of neck pain following mTBI was associated with PPCS at three months but not at twelve months (p=0.032 and p=0.584 respectively). Alcohol consumption around the time of injury was associated with PPCS at twelve months, however this should be interpreted with caution due to the relatively small number of participants who reported consuming alcohol (n=20) and wide confidence interval. LOC, retrograde and post-traumatic amnesia were not associated with PPCS at either timepoint. There was also no significant association between location of initial care (metropolitan vs regional hospitals) and PPCS at either three or twelve months

#### Site of head impact

The most frequently self-reported sites of head impact were to the right and left superior anterior aspects of the head (n=57 and 59 respectively) (Figure 5). Eight people (n=8) sustained a mTBI not related to head impact. A significant association was identified between impact to the left posterior superior aspect of the head and experiencing PPCS at three months (p=0.018) (Table 4). Those who experienced impact to the left anterior superior aspect of the head were more likely to experience PPCS at twelve months (p=0.046). Fewer people reported impact to the left posterior inferior aspect of their head (n=15 in total), but of those who completed twelve month follow up, 7/7 (100%) reported experiencing PPCS. Logistic regression could not be conducted for this relationship due to the small and imbalanced size in each recovery group. Notably, each of these significant associations were seen with impact to the left side of the head only. As participants could report impact to more than one region of their head, the relationship between PPCS and the number of regions affected was also investigated. There was no significant association between experiencing impact to multiple regions of the head and recovery at either three or twelve months.

**Figure 5.**
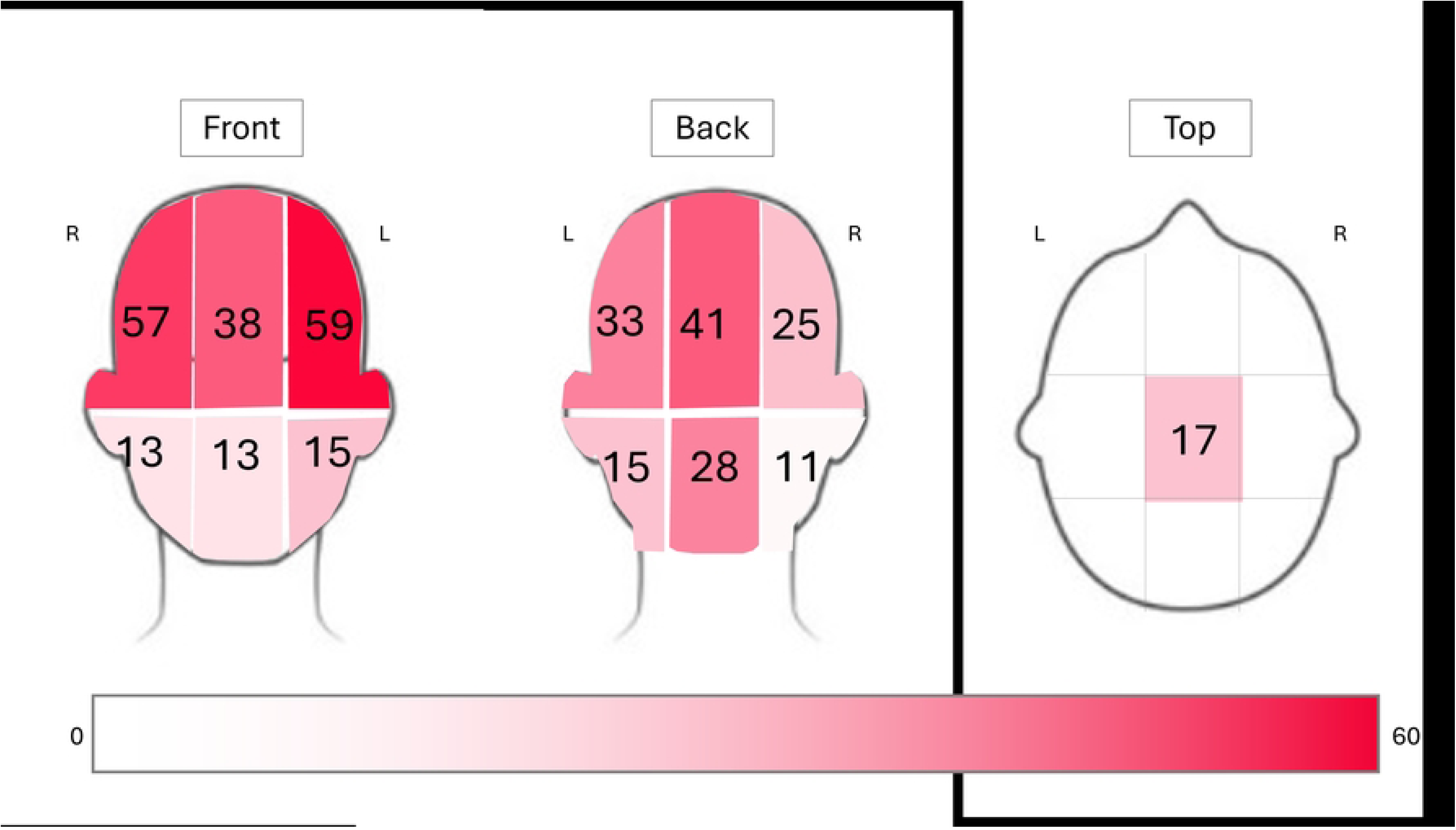
Shade and number represent overall number of participants who reported each segment as a site of impact for their mTBI. Participants could report more than one segment, and thus the total is greater than number of participants

**Table 4:** Univariate analysis of site of head impact as a predictor of PPCS at 3 and 12 months following mTBI.

|  | 3 months |  |  |  | 12 months |  |  |  |
| --- | --- | --- | --- | --- | --- | --- | --- | --- |
|  | No PPCS<br>(n=83)<br>n (%) | PPCS<br>(n=82)<br>n (%) | OR (95% CI) | p-value | No PPCS<br>(n=75)<br>n (%) | PPCS<br>(n=63)<br>n (%) | OR (95% CI) | p-value |
| **Site of head impact – individual** |  |  |  |  |  |  |  |  |
| Right anterior superior | 22 (26.5) | 21 (25.6) | 0.95 (0.47-1.91) | 0.895 | 22 (29.3) | 18 (28.6) | 0.96 (0.46-2.01) | 0.922 |
| Middle anterior superior | 11 (13.2) | 15 (18.3) | 1.46 (0.63-3.41) | 0.374 | 8 (10.7) | 12 (19.0) | 1.97 (0.75-5.17) | 0.164 |
| Left anterior superior | 17 (20.5) | 27 (32.9) | 1.90 (0.94-3.86) | 0.069 | 11 (14.7) | 18 (28.6) | 2.32 (1.00-5.39) | **0.046** |
| Right anterior inferior | 7 (8.4) | 5 (6.1) | 0.70 (0.21-2.31) | 0.562 | 4 (5.3) | 5 (7.9) | 1.53 (0.39-5.96) | 0.538 |
| Middle anterior inferior | 5 (6.0) | 6 (7.3) | 1.23 (0.36-4.20) | 0.739 | 6 (8.0) | 5 (7.9) | 0.99 (0.29-3.41) | 0.989 |
| Left anterior inferior | 8 (9.6) | 3 (3.7) | 0.36 (0.09-1.39) | 0.117 | 4 (5.3) | 5 (7.9) | 1.53 (0.39-5.96) | 0.538 |
| Left posterior superior | 6 (7.2) | 16 (19.5) | 3.11 (1.15-8.41) | **0.018** | 12 (16.0) | 8 (12.7) | 0.76 (0.29-2.00) | 0.582 |
| Middle posterior superior | 13 (15.7) | 14 (17.1) | 1.11 (0.48-2.53) | 0.806 | 13 (17.3) | 10 (15.9) | 0.89 (0.36-2.21) | 0.818 |
| Right posterior superior | 11 (13.2) | 9 (10.9) | 0.81 (0.31-2.06) | 0.654 | 10 (13.3) | 6 (9.5) | 0.68 (0.23-2.00) | 0.483 |
| Left posterior inferior | 4 (4.8) | 8 (9.8) | 2.13 (0.62-7.39) | 0.218 | 0 (0.0) | 7 (11.1) | (unable to conduct) * | **0.003** |
| Middle posterior inferior | 11 (13.2) | 11 (13.4) | 1.01 (0.41-2.49) | 0.976 | 13 (17.3) | 6 (9.5) | 0.50 (0.17-1.41) | 0.179 |
| Right posterior inferior | 3 (3.6) | 6 (7.3) | 2.10 (0.51-8.72) | 0.291 | 2 (2.7) | 6 (9.5) | 3.84 (0.75-19.75) | 0.082 |
| Top | 4 (4.8) | 7 (8.5) | 1.84 (0.51-6.55) | 0.336 | 6 (8.0) | 3 (4.8) | 0.57 (0.13-2.39) | 0.437 |
| **Site of head impact - multiple** |  |  |  | 0.201 |  |  |  | 0.404 |
| None | 4 (4.8) | 3 (3.7) | 0.96 (0.20-4.69) |  | 4 (5.3) | 1 (1.6) | 0.31 (0.03-2.95) |  |
| 1 | 46 (55.4) | 35 (42.7) | Reference |  | 39 (52.0) | 31 (49.2) | Reference |  |
| 2+ | 33 (39.8) | 44 (53.7) | 1.75 (0.93-3.29) |  | 32 (42.7) | 31 (49.2) | 1.21 (0.61-2.41) |  |

Number and percentage for No PPCS compared to PPCS at three and twelve months. Association between site of head impact and PPCS was assessed using logistic regression, with odds ratios and 95% confidence intervals reported. \*Logistic regression for the 'Left posterior inferior' region at 12 months could not be completed due to inadequate number of participants in the No PPCS group. Relationship between PPCS and those who experienced no direct head impact compared to those who had one site of head impact, or multiple sites of head impact are also reported.

Further exploratory analyses of the relationship between site of head impact and symptom presentation was conducted, based upon the hypothesis that impact to the posterior aspect of the head may affect function of the visual cortex. Of those who reported impact to the left posterior superior aspect of the head, 25/33 (75%) experienced light sensitivity at initial enrolment (p=0.010). Similarly, 13/15 (87%) who reported impact to the left posterior inferior aspect of their head experienced light sensitivity (p=0.011). Of those who reported impact to the left anterior superior aspect of their head (near the left eye), 32/58 (55%) experienced visual disturbances (p=0.001).

#### Relationship between initial symptom presentation and recovery

Exploratory analysis of symptoms at initial enrolment as predictors of PPCS at three and twelve months is provided in Table 5. Overall, initial symptom presentation was a strong predictor of PPCS at both three and twelve months. There was a significant difference in median number of symptoms, and symptom severity score, at initial presentation between those with PPCS compared to those without PPCS at both three and twelve months (all p<0.001). Specifically, participants who reported 12 or more symptoms initially were almost six times more likely to experience PPCS at three months (OR 5.94, 95% CI 3.03-11.63, p<0.001). Those with a symptom severity score of 30 or more had seven times the odds of poorer recovery (OR 7.26, 95% CI 3.63-14.49, p<0.001). Similar associations were seen for PPCS at twelve months (12 or more symptoms OR 5.58, 95% CI 2.68-11.61, p<0.001; symptom severity score of 30 or more OR 5.91, 95% CI 2.82-12.36, p<0.001).

**Table 5:**
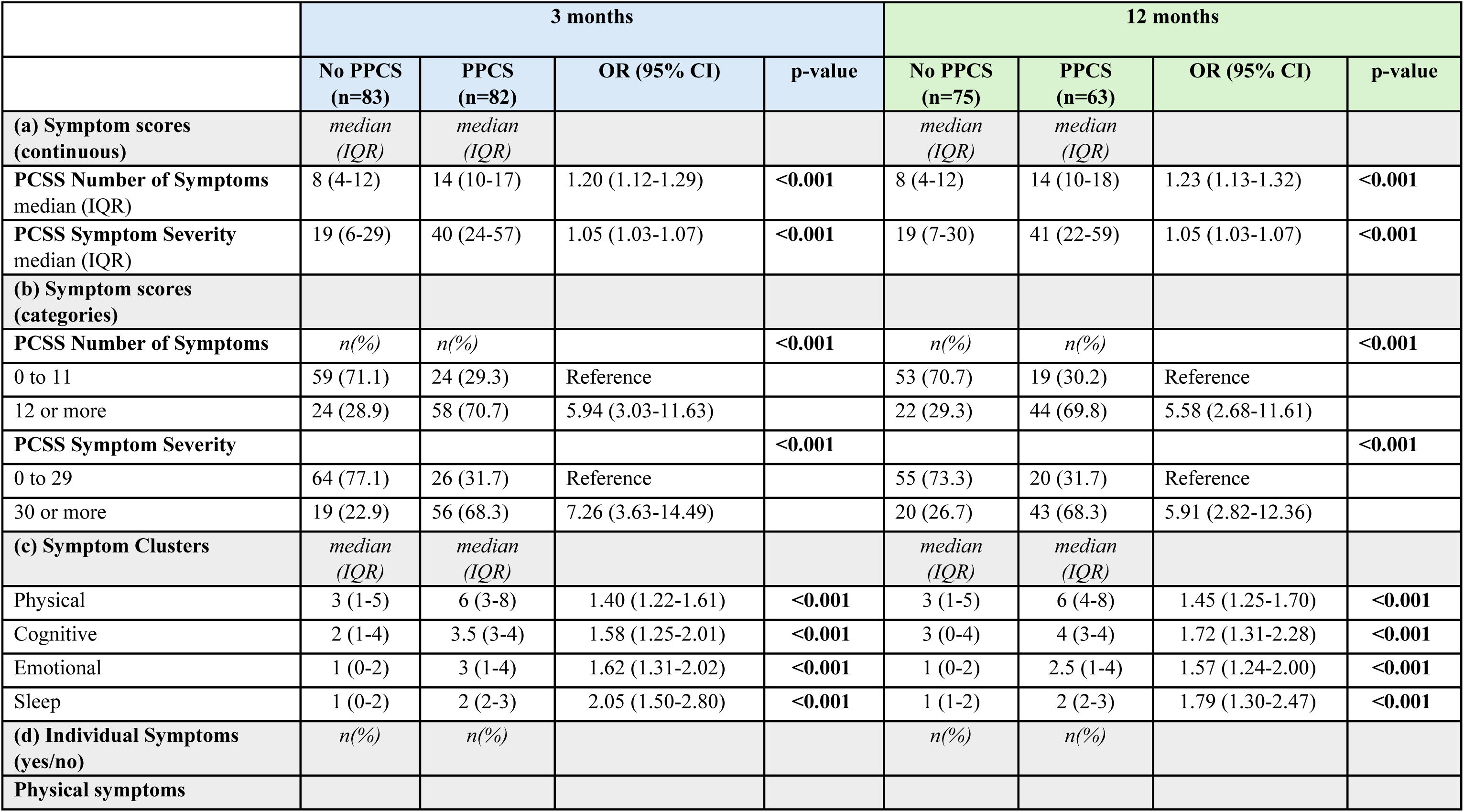
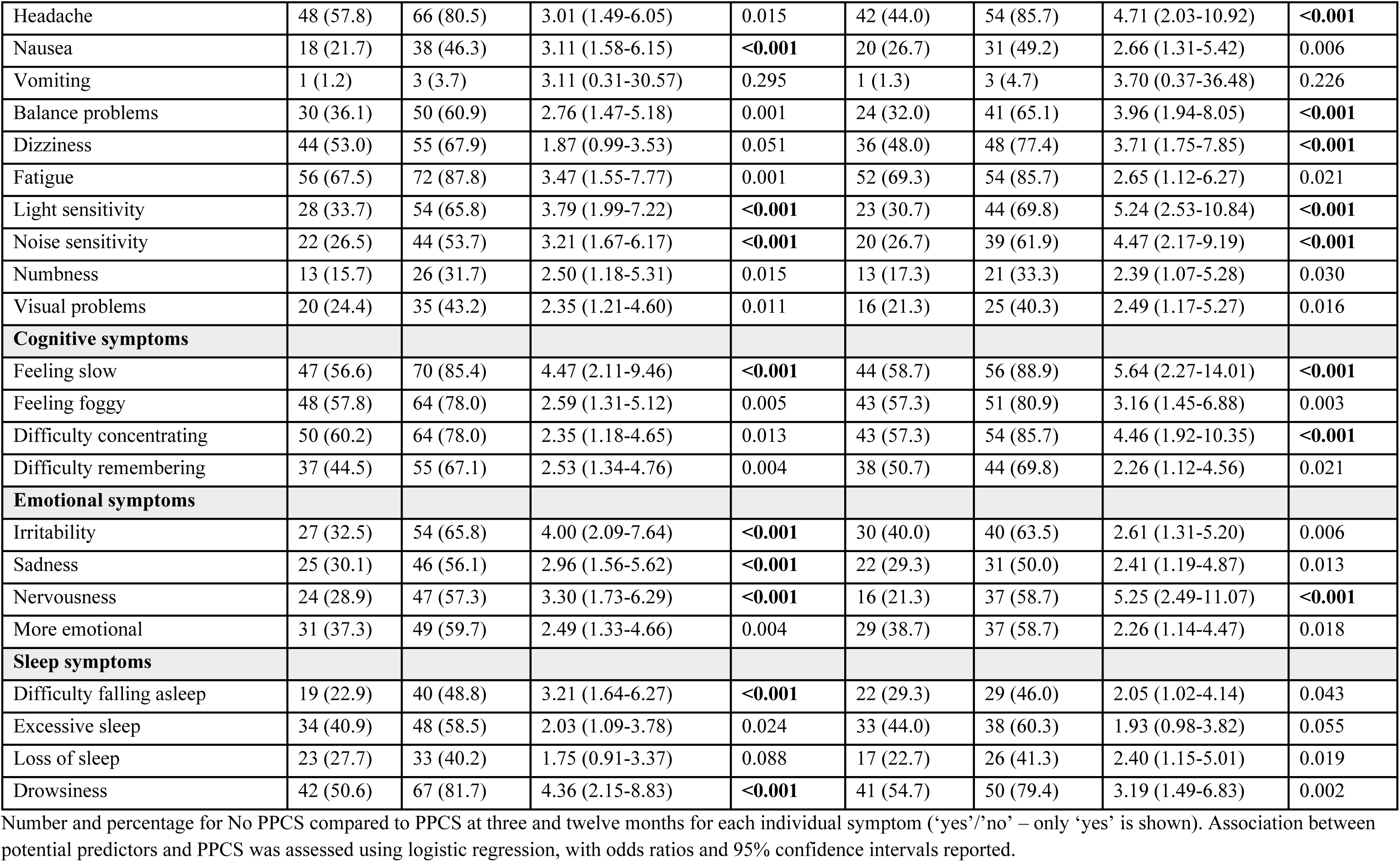
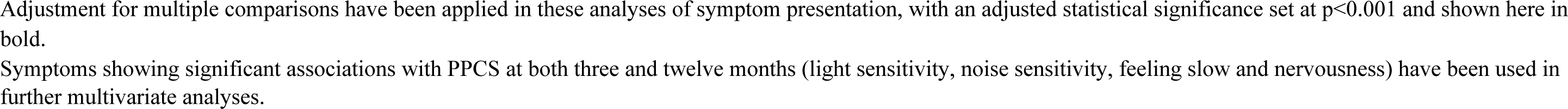
Exploratory analysis of symptoms at initial enrolment as predictors of outcome at 3 and 12 months following mTBI. Symptoms presented as (a) Symptom scores (continuous), (b) Symptom scores (categories using median cut-off), (c) Symptom Clusters (physical, cognitive, emotional and sleep) and (d) Individual Symptoms.

|  | 3 months |  |  |  | 12 months |  |  |  |
| --- | --- | --- | --- | --- | --- | --- | --- | --- |
|  | No PPCS<br>(n=83) | PPCS<br>(n=82) | OR (95% CI) | p-value | No PPCS<br>(n=75) | PPCS<br>(n=63) | OR (95% CI) | p-value |
| **(a) Symptom scores**<br>**(continuous)** | *median*<br>*(IQR)* | *median*<br>*(IQR)* |  |  | *median*<br>*(IQR)* | *median*<br>*(IQR)* |  |  |
| PCSS Number of Symptoms<br>median (IQR) | 8 (4-12) | 14 (10-17) | 1.20 (1.12-1.29) | <0.001 | 8 (4-12) | 14 (10-18) | 1.23 (1.13-1.32) | <0.001 |
| PCSS Symptom Severity<br>median (IQR) | 19 (6-29) | 40 (24-57) | 1.05 (1.03-1.07) | <0.001 | 19 (7-30) | 41 (22-59) | 1.05 (1.03-1.07) | <0.001 |
| **(b) Symptom scores**<br>**(categories)** |  |  |  |  |  |  |  |  |
| PCSS Number of Symptoms | *n(%)* | *n(%)* |  | <0.001 | *n(%)* | *n(%)* |  | <0.001 |
| 0 to 11 | 59 (71.1) | 24 (29.3) | Reference |  | 53 (70.7) | 19 (30.2) | Reference |  |
| 12 or more | 24 (28.9) | 58 (70.7) | 5.94 (3.03-11.63) |  | 22 (29.3) | 44 (69.8) | 5.58 (2.68-11.61) |  |
| PCSS Symptom Severity |  |  |  | <0.001 |  |  |  | <0.001 |
| 0 to 29 | 64 (77.1) | 26 (31.7) | Reference |  | 55 (73.3) | 20 (31.7) | Reference |  |
| 30 or more | 19 (22.9) | 56 (68.3) | 7.26 (3.63-14.49) |  | 20 (26.7) | 43 (68.3) | 5.91 (2.82-12.36) |  |
| **(c) Symptom Clusters** | *median*<br>*(IQR)* | *median*<br>*(IQR)* |  |  | *median*<br>*(IQR)* | *median*<br>*(IQR)* |  |  |
| Physical | 3 (1-5) | 6 (3-8) | 1.40 (1.22-1.61) | <0.001 | 3 (1-5) | 6 (4-8) | 1.45 (1.25-1.70) | <0.001 |
| Cognitive | 2 (1-4) | 3.5 (3-4) | 1.58 (1.25-2.01) | <0.001 | 3 (0-4) | 4 (3-4) | 1.72 (1.31-2.28) | <0.001 |
| Emotional | 1 (0-2) | 3 (1-4) | 1.62 (1.31-2.02) | <0.001 | 1 (0-2) | 2.5 (1-4) | 1.57 (1.24-2.00) | <0.001 |
| Sleep | 1 (0-2) | 2 (2-3) | 2.05 (1.50-2.80) | <0.001 | 1 (1-2) | 2 (2-3) | 1.79 (1.30-2.47) | <0.001 |
| **(d) Individual Symptoms**<br>**(yes/no)** | *n(%)* | *n(%)* |  |  | *n(%)* | *n(%)* |  |  |
| Physical symptoms |  |  |  |  |  |  |  |  |
| Headache | 48 (57.8) | 66 (80.5) | 3.01 (1.49-6.05) | 0.015 | 42 (44.0) | 54 (85.7) | 4.71 (2.03-10.92) | <0.001 |
| Nausea | 18 (21.7) | 38 (46.3) | 3.11 (1.58-6.15) | <0.001 | 20 (26.7) | 31 (49.2) | 2.66 (1.31-5.42) | 0.006 |
| Vomiting | 1 (1.2) | 3 (3.7) | 3.11 (0.31-30.57) | 0.295 | 1 (1.3) | 3 (4.7) | 3.70 (0.37-36.48) | 0.226 |
| Balance problems | 30 (36.1) | 50 (60.9) | 2.76 (1.47-5.18) | 0.001 | 24 (32.0) | 41 (65.1) | 3.96 (1.94-8.05) | <0.001 |
| Dizziness | 44 (53.0) | 55 (67.9) | 1.87 (0.99-3.53) | 0.051 | 36 (48.0) | 48 (77.4) | 3.71 (1.75-7.85) | <0.001 |
| Fatigue | 56 (67.5) | 72 (87.8) | 3.47 (1.55-7.77) | 0.001 | 52 (69.3) | 54 (85.7) | 2.65 (1.12-6.27) | 0.021 |
| Light sensitivity | 28 (33.7) | 54 (65.8) | 3.79 (1.99-7.22) | <0.001 | 23 (30.7) | 44 (69.8) | 5.24 (2.53-10.84) | <0.001 |
| Noise sensitivity | 22 (26.5) | 44 (53.7) | 3.21 (1.67-6.17) | <0.001 | 20 (26.7) | 39 (61.9) | 4.47 (2.17-9.19) | <0.001 |
| Numbness | 13 (15.7) | 26 (31.7) | 2.50 (1.18-5.31) | 0.015 | 13 (17.3) | 21 (33.3) | 2.39 (1.07-5.28) | 0.030 |
| Visual problems | 20 (24.4) | 35 (43.2) | 2.35 (1.21-4.60) | 0.011 | 16 (21.3) | 25 (40.3) | 2.49 (1.17-5.27) | 0.016 |
| **Cognitive symptoms** |  |  |  |  |  |  |  |  |
| Feeling slow | 47 (56.6) | 70 (85.4) | 4.47 (2.11-9.46) | <0.001 | 44 (58.7) | 56 (88.9) | 5.64 (2.27-14.01) | <0.001 |
| Feeling foggy | 48 (57.8) | 64 (78.0) | 2.59 (1.31-5.12) | 0.005 | 43 (57.3) | 51 (80.9) | 3.16 (1.45-6.88) | 0.003 |
| Difficulty concentrating | 50 (60.2) | 64 (78.0) | 2.35 (1.18-4.65) | 0.013 | 43 (57.3) | 54 (85.7) | 4.46 (1.92-10.35) | <0.001 |
| Difficulty remembering | 37 (44.5) | 55 (67.1) | 2.53 (1.34-4.76) | 0.004 | 38 (50.7) | 44 (69.8) | 2.26 (1.12-4.56) | 0.021 |
| **Emotional symptoms** |  |  |  |  |  |  |  |  |
| Irritability | 27 (32.5) | 54 (65.8) | 4.00 (2.09-7.64) | <0.001 | 30 (40.0) | 40 (63.5) | 2.61 (1.31-5.20) | 0.006 |
| Sadness | 25 (30.1) | 46 (56.1) | 2.96 (1.56-5.62) | <0.001 | 22 (29.3) | 31 (50.0) | 2.41 (1.19-4.87) | 0.013 |
| Nervousness | 24 (28.9) | 47 (57.3) | 3.30 (1.73-6.29) | <0.001 | 16 (21.3) | 37 (58.7) | 5.25 (2.49-11.07) | <0.001 |
| More emotional | 31 (37.3) | 49 (59.7) | 2.49 (1.33-4.66) | 0.004 | 29 (38.7) | 37 (58.7) | 2.26 (1.14-4.47) | 0.018 |
| **Sleep symptoms** |  |  |  |  |  |  |  |  |
| Difficulty falling asleep | 19 (22.9) | 40 (48.8) | 3.21 (1.64-6.27) | <0.001 | 22 (29.3) | 29 (46.0) | 2.05 (1.02-4.14) | 0.043 |
| Excessive sleep | 34 (40.9) | 48 (58.5) | 2.03 (1.09-3.78) | 0.024 | 33 (44.0) | 38 (60.3) | 1.93 (0.98-3.82) | 0.055 |
| Loss of sleep | 23 (27.7) | 33 (40.2) | 1.75 (0.91-3.37) | 0.088 | 17 (22.7) | 26 (41.3) | 2.40 (1.15-5.01) | 0.019 |
| Drowsiness | 42 (50.6) | 67 (81.7) | 4.36 (2.15-8.83) | <0.001 | 41 (54.7) | 50 (79.4) | 3.19 (1.49-6.83) | 0.002 |

Number and percentage for No PPCS compared to PPCS at three and twelve months for each individual symptom ('yes'/'no' – only 'yes' is shown). Association between potential predictors and PPCS was assessed using logistic regression, with odds ratios and 95% confidence intervals reported.
Adjustment for multiple comparisons have been applied in these analyses of symptom presentation, with an adjusted statistical significance set at $p < 0.001$ and shown here in bold. Symptoms showing significant associations with PPCS at both three and twelve months (light sensitivity, noise sensitivity, feeling slow and nervousness) have been used in further multivariate analyses.

The relationship between commonly recognised symptom clusters (Pardini, 2004) and PPCS was investigated. Each cluster (physical, cognitive, emotional and sleep) was associated with PPCS at both three and twelve months (Table 5). However, in some cases the difference in median scores between those with No PPCS compared to those with PPCS was small and may not represent a clinically significant difference. The largest differences in median scores for those with no PPCS compared to those with PPCS was seen between the physical (headache, nausea, vomiting, balance, dizziness, visual problems, fatigue, light sensitivity, noise sensitivity, numbness) and emotional (irritable, sadness, more emotional, nervousness) clusters. For each additional physical symptom reported at initial enrolment there was 1.40 odds of experiencing PPCS at three months (OR 1.40, 95%CI 1.22-1.61, p,0.001). For each additional emotional symptom, the odds of experiencing PPCS at three months was 1.6 times (OR 1.62, 95% CI 1.31-2.02, p<0.001) greater.

Each individual symptom at initial enrolment was also assessed for association with PPCS at three and twelve months. Several symptoms were associated with PPCS at each timepoint (all p<0.001, see Table 5 for details of specific symptoms and odds ratios). Those with more emotional symptoms (irritability, sadness and nervousness) were more likely to experience PPCS at three months, whilst physical symptoms (headache, balance problems, dizziness, light and noise sensitivity) were associated with PPCS at twelve months. Of note, light and noise sensitivity, feeling slow and nervousness were associated with PPCS at both timepoints.

Participants presenting with those symptoms were three to five times more likely to experience PPCS compared to those who did not report those symptoms.

### Multivariable modelling of PPCS

Table 6 provides an overview of statistical findings for each model examined, adjusted for all other variables, at both three and twelve months. As symptom presentation at initial enrolment appeared to be the most consistent predictor of PPCS, we chose to investigate several variations of symptom presentation as primary predictors in multivariable modelling.

**Table 6:**
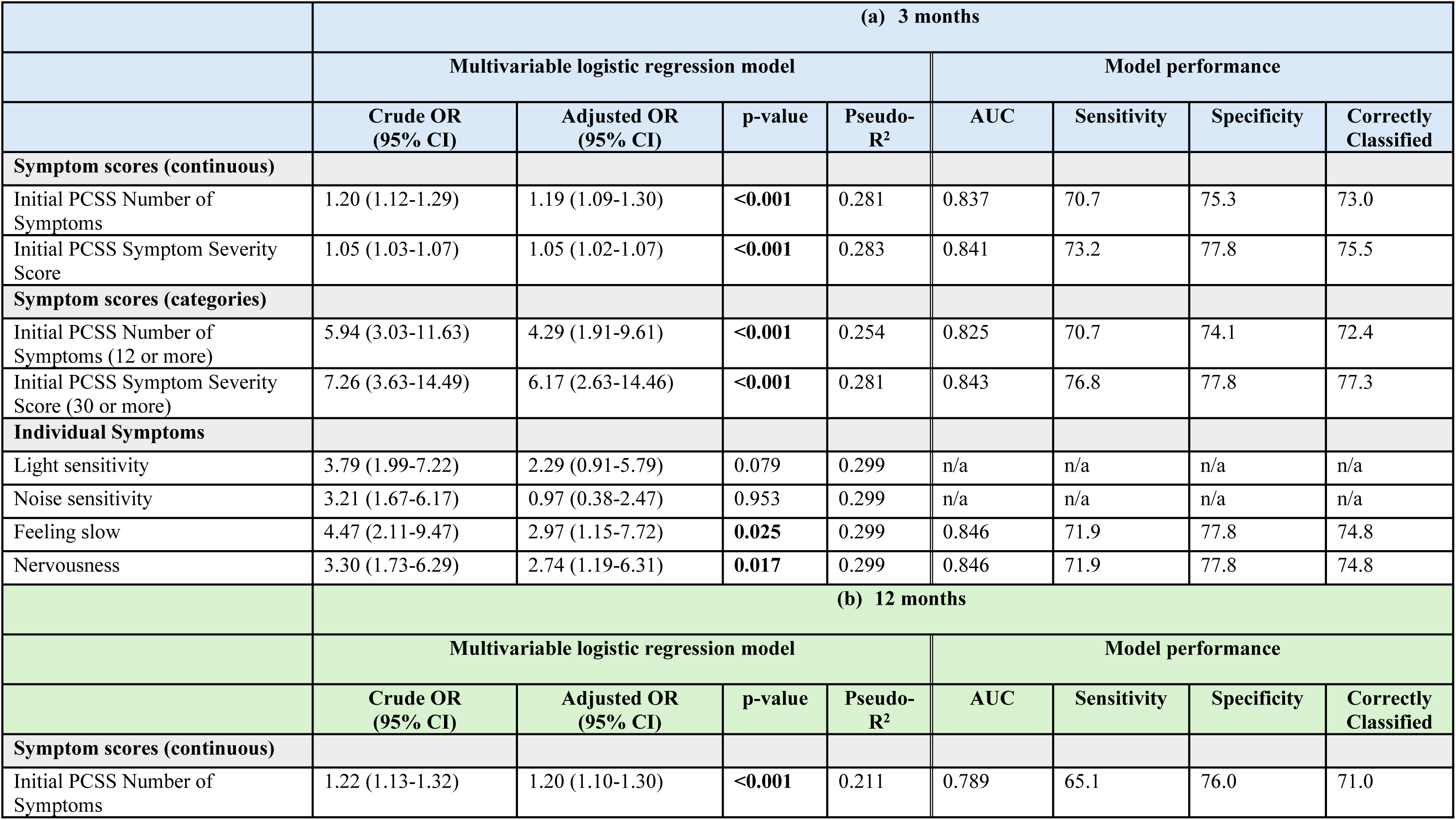
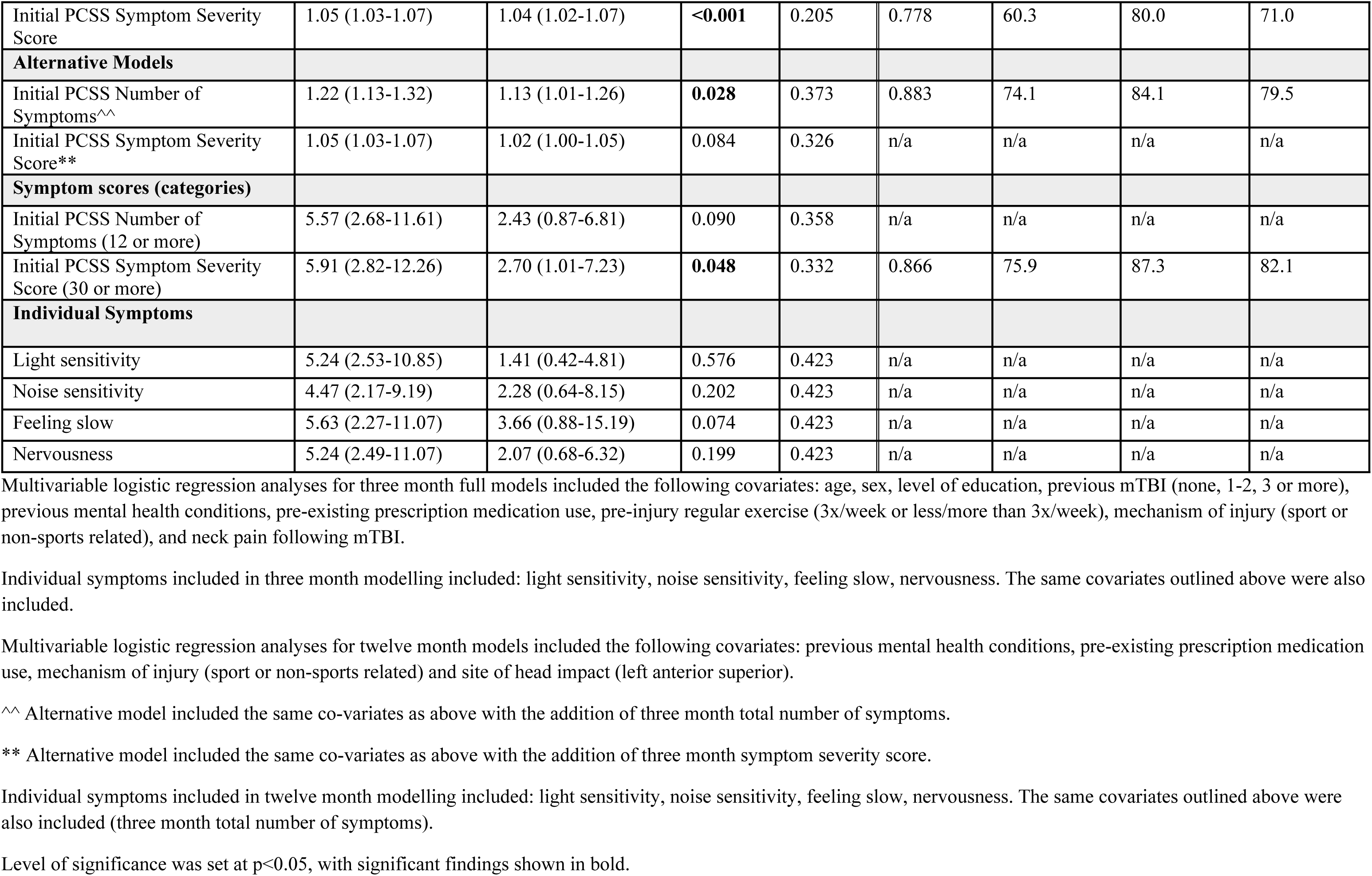
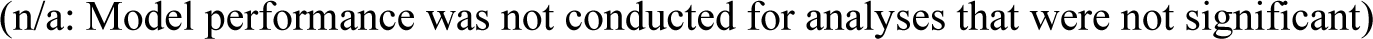
Association between primary variables of interest and PPCS using multivariable logistic regression modelling, and model performance, at (a) 3 months and (b) 12 months.

|  | (a) 3 months |  |  |  |  |  |  |  |
| --- | --- | --- | --- | --- | --- | --- | --- | --- |
|  | Multivariable logistic regression model |  |  |  | Model performance |  |  |  |
|  | Crude OR<br>(95% CI) | Adjusted OR<br>(95% CI) | p-value | Pseudo-<br>R<sup>2</sup> | AUC | Sensitivity | Specificity | Correctly<br>Classified |
| **Symptom scores (continuous)** |  |  |  |  |  |  |  |  |
| Initial PCSS Number of Symptoms | 1.20 (1.12-1.29) | 1.19 (1.09-1.30) | <0.001 | 0.281 | 0.837 | 70.7 | 75.3 | 73.0 |
| Initial PCSS Symptom Severity Score | 1.05 (1.03-1.07) | 1.05 (1.02-1.07) | <0.001 | 0.283 | 0.841 | 73.2 | 77.8 | 75.5 |
| **Symptom scores (categories)** |  |  |  |  |  |  |  |  |
| Initial PCSS Number of Symptoms (12 or more) | 5.94 (3.03-11.63) | 4.29 (1.91-9.61) | <0.001 | 0.254 | 0.825 | 70.7 | 74.1 | 72.4 |
| Initial PCSS Symptom Severity Score (30 or more) | 7.26 (3.63-14.49) | 6.17 (2.63-14.46) | <0.001 | 0.281 | 0.843 | 76.8 | 77.8 | 77.3 |
| **Individual Symptoms** |  |  |  |  |  |  |  |  |
| Light sensitivity | 3.79 (1.99-7.22) | 2.29 (0.91-5.79) | 0.079 | 0.299 | n/a | n/a | n/a | n/a |
| Noise sensitivity | 3.21 (1.67-6.17) | 0.97 (0.38-2.47) | 0.953 | 0.299 | n/a | n/a | n/a | n/a |
| Feeling slow | 4.47 (2.11-9.47) | 2.97 (1.15-7.72) | 0.025 | 0.299 | 0.846 | 71.9 | 77.8 | 74.8 |
| Nervousness | 3.30 (1.73-6.29) | 2.74 (1.19-6.31) | 0.017 | 0.299 | 0.846 | 71.9 | 77.8 | 74.8 |
|  | (b) 12 months |  |  |  |  |  |  |  |
|  | Multivariable logistic regression model |  |  |  | Model performance |  |  |  |
|  | Crude OR<br>(95% CI) | Adjusted OR<br>(95% CI) | p-value | Pseudo-<br>R<sup>2</sup> | AUC | Sensitivity | Specificity | Correctly<br>Classified |
| **Symptom scores (continuous)** |  |  |  |  |  |  |  |  |
| Initial PCSS Number of Symptoms | 1.22 (1.13-1.32) | 1.20 (1.10-1.30) | <0.001 | 0.211 | 0.789 | 65.1 | 76.0 | 71.0 |
| Initial PCSS Symptom Severity Score | 1.05 (1.03-1.07) | 1.04 (1.02-1.07) | **<0.001** | 0.205 | 0.778 | 60.3 | 80.0 | 71.0 |
| **Alternative Models** |  |  |  |  |  |  |  |  |
| Initial PCSS Number of Symptoms^^ | 1.22 (1.13-1.32) | 1.13 (1.01-1.26) | **0.028** | 0.373 | 0.883 | 74.1 | 84.1 | 79.5 |
| Initial PCSS Symptom Severity Score** | 1.05 (1.03-1.07) | 1.02 (1.00-1.05) | 0.084 | 0.326 | n/a | n/a | n/a | n/a |
| **Symptom scores (categories)** |  |  |  |  |  |  |  |  |
| Initial PCSS Number of Symptoms (12 or more) | 5.57 (2.68-11.61) | 2.43 (0.87-6.81) | 0.090 | 0.358 | n/a | n/a | n/a | n/a |
| Initial PCSS Symptom Severity Score (30 or more) | 5.91 (2.82-12.26) | 2.70 (1.01-7.23) | **0.048** | 0.332 | 0.866 | 75.9 | 87.3 | 82.1 |
| **Individual Symptoms** |  |  |  |  |  |  |  |  |
| Light sensitivity | 5.24 (2.53-10.85) | 1.41 (0.42-4.81) | 0.576 | 0.423 | n/a | n/a | n/a | n/a |
| Noise sensitivity | 4.47 (2.17-9.19) | 2.28 (0.64-8.15) | 0.202 | 0.423 | n/a | n/a | n/a | n/a |
| Feeling slow | 5.63 (2.27-11.07) | 3.66 (0.88-15.19) | 0.074 | 0.423 | n/a | n/a | n/a | n/a |
| Nervousness | 5.24 (2.49-11.07) | 2.07 (0.68-6.32) | 0.199 | 0.423 | n/a | n/a | n/a | n/a |

(n/a: Model performance was not conducted for analyses that were not significant)

#### Predictive models of outcome at three months

Three-month models were adjusted for all other predictors that were statistically significant on univariate analyses (age, sex, level of education, previous mTBI (none, 1-2, 3 or more), previous mental health conditions, pre-existing prescription medication use, pre-injury regular exercise, mechanism of injury (sport or non-sports related) and neck pain following mTBI). The primary predictor for the first models used symptom scores (number and severity) as continuous variables. For the Number of Symptoms there was an adjusted OR of 1.19 (95%CI 1.09-1.30, p<0.001, pseudo-R^2^=0.281), indicating that for each additional symptom there was a 19% increase in odds of PPCS. For each additional increase in symptom severity score there was a 5% increase in odds of PPCS (aOR = 1.05, 95%CI 1.02-1.07, p<0.001, pseudo-R^2^=0.283).

In an effort to improve applicability and interpretability in a clinical setting, binarised variables and individual symptoms as primary predictors were investigated. For the binary variables, the best performing model indicated that participants with a ‘high’ initial PCSS Symptom Severity Score (30 or more) were six times more likely to experience PPCS at three months (aOR=6.17, 95%CI =2.63-14.46, p<0.001, pseudo-R^2^=0.281), after adjusting for age, sex, level of education, previous mTBI (none, 1-2, 3 or more), previous mental health conditions, pre-existing prescription medication use, pre-injury regular exercise (more than 3x/week), mechanism of injury (sport/non-sports related), and neck pain following injury. For this model, area under the curve (AUC) was 0.843, sensitivity was 76.8%, specificity 77.8%, with 77.3% of cases correctly classified. Of these co-variates, previous mental health conditions was also a strong predictor within the model (p=0.001).

Acute individual symptoms including feeling slow and nervousness were also associated with PPCS at three months on multivariate analysis (aOR=2.97, 95%CI 1.15-7.72, p= 0.025, pseudo-R^2^=0.299; and aOR=2.74, 95%CI 1.19-6.31, p=0.017, pseudo-R^2^=0.299 respectively). For these models, AUC was 0.846, sensitivity was 71.9%, specificity 77.8%, with 74.8% of cases correctly classified (Table 6a).

#### Predictive models of outcome at twelve months

Twelve-month models were adjusted for all other predictors that were statistically significant on univariate analyses, including previous mental health conditions, pre-existing prescription medication use, mechanism of injury (sport or non-sports related) and location of head impact (left anterior superior aspect). Symptom scores were first investigated as continuous variables: for the primary predictor Number of Symptoms there was an adjusted OR of 1.20 (95%CI 1.10-1.30, p<0.001, pseudo-R^2^=0.211), indicating that for each additional symptom reported there was a 20% increase in odds of PPCS. For the model with Symptom Severity Score as primary predictor, there was a 4% increase in odds of PPCS for each additional point increase in symptom severity score (aOR = 1.04, 95%CI 1.02-1.07, p<0.001, pseudo-R^2^=0.205). There was a notable increase in model performance when three month symptom scores were also included in the model as a co-variate for the model using ‘Number of Symptoms’ (aOR 1.13, 95%CI 1.01-1.26, p=0.028, pseudo-R^2^=0.373), but the model using Symptom Severity Score was no longer significant (aOR 1.02, 95%CI 1.00-1.05, p=0.084, pseudo-R^2^=0.326).

For the binary variables, the best performing model (adjusted for previous mental health conditions, pre-existing prescription medication use, mechanism of injury, site of head impact and three month PCSS number of symptoms) indicated that participants with a ‘high’ initial PCSS Symptom Severity Score (30 or more) were almost three times more likely to experience PPCS at twelve months (aOR=2.70, 95%CI =1.01-7.23, p=0.048, pseudo-R^2^=0.332). For this model, area under the curve (AUC) was 0.866, sensitivity was 75.9%, specificity 87.3%, with 82.1% of cases correctly classified (Table 6b). No significant associations were found between acute individual symptoms (light or noise sensitivity, feeling slow and nervousness) and PPCS at twelve months on multivariate analysis.

## Discussion

This study provides valuable insights into recovery after mTBI in a community-based adult cohort and identifies predictive factors for people at risk of developing PPCS at three and twelve months post-injury. Of those participants who completed follow-up, almost half were still experiencing persisting symptoms three and twelve months after mTBI. PPCS was associated with lower quality of life and a delay resuming normal physical activity but was not associated with returning to work or study at three and twelve months. After considering the effect of biological factors (such as age, sex) and other demographic and lifestyle factors, symptom burden (expressed as either number or total severity of symptoms using the Post-Concussion Symptom Scale) was found to be the strongest predictor of PPCS at both three and twelve months.

### Recovery outcomes over time

The number of people in this study who experienced PPCS in the first twelve months post-mTBI is consistent with previous literature examining mTBI cohorts in a non-athletic population(46), but is higher than the 31.3% prevalence of PPCS reported in a recent meta-analysis that included studies of all-cause mTBI(8). There was variability in recovery trajectories, indicated by the presence or absence of PPCS across time points. This underscores the unpredictable nature of mTBI and the need for ongoing monitoring and individualised support for people with persisting symptoms after mTBI(9).

### Relationship between functional recovery, quality of life and PPCS outcomes

To our knowledge, this is the first study to report that many people who had returned to work were still experiencing symptoms at three or twelve months post mTBI. Similar to other studies, nearly all participants who were working prior to their mTBI had returned to work by three months(14), however more than 40% continued to report experiencing PPCS. Returning to work despite having PPCS may reflect socioeconomic pressures, with the potential for financial stress as a result of having time off work. Individuals may implement workplace accommodations such as reduced hours, flexible work arrangements or a change in position, to help to facilitate an earlier RTW after mild TBI(17) though this information was not collected as part of this study. Even with accommodations, workers with persisting symptoms may be more likely to have reduced productivity at work(16,47), limitations in their work capacity(16) and be at greater risk of re-injury(48). Employers may benefit from specific guidance on accommodations to facilitate RTW, ensuring they have an understanding on how PPCS may impact an individual’s work performance as they recover. Further research into the continuum of RTW after mTBI is necessary to understand the socioeconomic cost and burden of PPCS on an individual’s physical and emotional well-being and sense of identity.

In contrast to RTW, individuals with persisting symptoms were significantly less likely to have returned to normal physical activity post-injury, possibly suggesting a prioritisation of vocational return over physical recovery. People may also avoid or delay returning to physical activity because of exercise intolerance, fear of worsening symptoms or concern about sustaining another mTBI. However, recent evidence supports the beneficial nature of exercise on recovery(49–52), and it is possible that early return to physical activity may reduce the risk of experiencing PPCS. In support of this, we found that individuals who engaged in regular physical activity prior to injury (more than three times per week) were less likely to develop PPCS. This may reflect the protective influence of higher baseline physical fitness, as well as a greater tendency to re-establish pre-injury routines earlier in recovery. Meeting physical activity guidelines post-mTBI is also important to reduce the impact of PPCS on quality of life, fatigue, cognition and adverse mental health outcomes(15). Healthcare providers should prioritise patient education on the importance of a safe, early return to physical activity, along with practical guidance to support and facilitate this process for people with mTBI soon after injury.

### Relationship between predictive factors of interest and PPCS

The results of this study indicate that initial symptom burden, expressed as either number of symptoms, total severity of symptoms, or individual symptoms were the strongest predictors of PPCS at both three and twelve months. This result aligns with findings from a recent systematic review which reported that the most commonly reported prognostic factors were baseline symptom severity (individual symptoms and validated symptom scales)(53). Using symptom presentation as a predictor of recovery is advantageous in clinical settings, with symptom reports a key component of standard clinical assessment. Here, we have modelled symptom presentation using various classification methods in an effort to reflect the various ways symptom assessments can be implemented in a clinical setting.

Whilst validated symptom scales are widely used in a research capacity, they are less commonly used by acute care clinicians(54,55). Anecdotally, clinicians report asking patients to self-select the symptoms they are experiencing, or alternatively, ask them about individual symptoms commonly experienced following mTBI (for example, headaches or dizziness). We found that, after adjusting for co-variates, specific symptoms including ‘feeling slow’ and ‘nervousness’ were predictive of PPCS at three months. Answering questions slowly has previously been identified as a predictor of persistent symptoms in children(56), and may reflect impairments in neurotransmission and cognitive processing related to injury pathophysiology(57). ‘Feeling nervous’ could plausibly be considered a proxy for anxiety and is an important symptom presentation to consider with anxiety (both pre-existing and post-injury) consistently associated with poorer outcomes following mTBI(53,58,59).

Our finding that model performance improved by including symptom presentation at three months in a multivariable model of PPCS at twelve months suggests that initial prognostic factors may be less influential with time. This is indicated by stronger explanatory power in the model when including three month symptom burden (R^2^ = 0.152 vs 0.305), highlighting the importance of ongoing symptom assessment to identify individuals at risk of longer-term symptoms and refer for necessary healthcare input accordingly.

One of the main limitations to symptoms as a prognostic factor is the possibility of under-reporting (for example, by athletes wanting to return to play)(60), or over-reporting (for example, by those likely to benefit from secondary gain)(61). Thus, there is an ongoing need to investigate and identify objective prognostic indicators. A comprehensive approach incorporating clinical variables, biomarkers (blood and saliva)(62,63), imaging (CT and MRI pathological observations), and other modifying factors has recently described in a CBI-M framework(64), and may play a role in predicting mTBI recovery.

#### Demographic factors

Whilst extensively studied, the relationship between demographic factors and recovery continues to show mixed results in the literature(53). We identified that older age, female sex and level of education were individually associated with PPCS at three-months post injury however these were not predictive of outcome at twelve months. Previous literature has identified that females(65) and older adults(65) experience more gradual symptom resolution and delayed recovery trajectories, and the current data support these findings, highlighting the need to consider the effects of these variables in comprehensive prediction modelling.

#### Pre-Injury Factors

Pre-existing mental health issues, sleep disorders and regular prescription medication use were each significantly associated with PPCS at three and twelve months, consistent with previous research highlighting the prognostic importance of pre-morbid health status in recovery after mTBI(23,27). Pre-existing mental health conditions such as depression, anxiety or post-traumatic stress disorder are amongst the strongest and most consistent predictors of delayed recovery after mTBI(7,21,23,27). As a result of such conditions, these individuals may have altered stress responses, reduced resilience to injury, or heightened symptom perception, all of which may contribute to prolonged symptom duration(66). Similarly, pre-injury sleep disorders were also associated with increased odds of PPCS at both time points, reflecting the growing evidence that disrupted sleep before or after injury can negatively influence recovery trajectories(67). Poor sleep has been linked to impaired neuroplasticity, reduced glymphatic clearance, impaired emotional regulation, and greater fatigue(68,69) which may prolong PPCS. These results emphasise the need for post-injury support for individuals with known pre-existing mental health or sleep problems to mitigate the risk of long-term PPCS following mTBI.

Individuals who reported three or more previous mTBIs were more likely to have PPCS at three months, consistent with previous literature(22). Interestingly, in this study we found that those who had experienced 1-2 prior mTBI’s were *less* likely to experience PPCS at three months compared to no previous mTBI. It may be hypothesised that people who have experienced 1-2 prior mTBIs experienced these in a sport-related context, and thus may be more familiar with likely outcomes and the return to physical activity process, thus reducing the likelihood of developing PPCS. They may also have a higher level of pre-injury fitness and better lifestyle routines around physical activity. However, given the widespread media attention on chronic traumatic encephalopathy in recent times, those who have had repeated mTBI’s may experience some anxiety around the long term consequences of these injuries, and potentially impact recovery. Further exploration of this association will provide better understanding of how prior injury may impact the development of PPCS.

#### Peri-injury factors

Our finding that people who experienced mTBI from non-sport related mechanisms were more likely to experience PPCS is consistent with other literature(53). People who experience mTBI in a sporting context will likely have better access to healthcare management and post-injury protocols(53), and may also have better general health and less likelihood of comorbidities(70). Indeed, our finding of an association between prescription medication use prior to injury, as a proxy of co-morbidities, and PPCS at three months supports this concept. Non-sport related mTBI, particularly those involving traumatic events such as transport accidents or assaults, may involve an overlay of acute psychological stress that impacts a person’s mental health and thus, their likelihood of a timely recovery(71)

Loss of consciousness and post-traumatic amnesia at the time of the injury were not found to be significant predictors of PPCS in this study, contributing to the evidence that perceived injury severity is not associated with the development of persisting symptoms or poorer functional recovery(72–74). In a clinical context, this may have implications for the diagnosis of mTBI and rather than the presence of a brief period of LOC or PTA indicating a *mild* TBI, it may be more accurate that these clinical factors are more applicable to exclude more severe injury. Particularly when considered in conjunction with previous research demonstrating a lack of association between perceived injury severity and poorer recovery(73), these findings challenge the concept that PPCS is a direct consequence of neurobiological injury to the brain. Providing education and follow-up care to individuals with clinically less severe mTBI may help reduce the burden of PPCS.

#### Site of head impact

Literature on the relationship between location of head impact and mTBI response is scant, though one study found no association between head biomechanics or impact location and symptom burden or recovery^57^. Our study used a novel approach to capture the location of head impact during the mTBI incident and identified several findings of interest between site of impact and recovery. Notably, each of the significant findings involved impact to the left side of the head. This apparent lateralisation effect may be explained by the fact that more of the population are right-side dominant, and impact to the left side of the head would predominantly affect right-sided function. Theoretically, it could be expected that different impact locations would produce tissue injury in different brain regions, with resultant functional impairments specific to that region(75). This is supported by the exploratory analyses in this study that identified several associations of interest between impact to the posterior aspect of the head and visual disturbances and light sensitivity. Although this was the not the primary objective for this study, it alludes to the need for further exploration of relationships between site of head impact and symptom presentation. Emerging sensor-based technologies used in a sporting context may provide objective data regarding head impact location and kinematics may have a role in further assessing this relationship.

#### Clinical Applications

To usefully predict PPCS it is necessary to identify models with an optimal balance of sensitivity and specificity. From a clinical perspective, higher sensitivity scores are preferrable to support the correct early identification of those who are likely to develop PPCS and direct them to appropriate treatment as required. Higher specificity (that is, the ability to correctly identify those who are *not* likely to develop PPCS), is of less concern clinically as it would not be deleterious if patients were referred for further follow-up but did not subsequently develop PPCS. However, there may be potential economic consequences due to unnecessary treatment and potential cost to patients depending upon the healthcare setting. It is important to note that care should be exercised when discussing prognostic outcomes with patients due to the potential for unintended psychological overlay leading to worsening symptoms if people are told they are higher risk for potential poorer recovery(76,77). The moderate-high sensitivity and specificity of our best performing models should give clinicians confidence in using symptom severity, or the individual symptoms of ‘feeling slow’ and ‘nervousness’, to predict those at risk or prolonged recovery. Regular ongoing monitoring of people who present with high symptom severity should be included in the care pathway for people after mTBI, including referral to specialist rehabilitation services if symptoms persist.

### Strengths

This study fills a crucial gap in the literature by focusing on the impact of mTBI on individuals in the general population who have experienced mTBI from a variety of causes, with consideration to symptom burden, quality of life and return to functional activities. This reflects the heterogeneity of mTBI. Our findings complement and augment prior work through the inclusion of a broad array of predictive factors in multivariable modelling, and an extensive longitudinal follow-up to twelve months. The nature of data collection involved in this study also enabled participant recruitment from multiple hospital sites and clinics, across metropolitan and regional areas, adding to the generalisability of the study findings. Additionally, this work provides a novel concept for assessing the site of head impact which may guide future research directions to further test and develop this concept.

There is a need for effective actions that can be applied to a clinical setting to improve practice guidelines(37). A further strength of this work is the clinical lens that has been applied to the study design, statistical analysis and reporting of results. This work will enable healthcare professionals to more easily identify people at greatest risk of PPCS and ensure timely and personalised follow up care is able to be provided.

### Limitations

Within the mTBI literature there is currently a lack of consensus on how recovery, or persisting symptoms, should be defined and measured(78). Most studies use self-reported symptoms, although the criteria for cut-off scores are not well defined. For the PCSS, symptom recovery has been defined in some studies by a symptom score of zero, others have used ‘return to ‘baseline’, and others have used a cut-off score of less than seven as defined in this study(78,79). Furthermore, the PCSS does not account for pre-existing symptoms that may present in the general population, as symptoms associated with mTBI are non-specific and can be present in many other medical conditions(80). Recent systematic reviews of criteria used to define recovery from sport-related concussion suggest that using a combination of symptom reports in conjunction with validated physiological measures and clinical examination would provide a more objective and reproducible definition of recovery(79,81). Unfortunately, this was not possible in the current study as follow-ups were conducted by telephone.

#### Selection bias

As the majority of participants in this study were referred via Emergency Departments, they may have tended to be more ‘significant’ mTBI and this factor should be considered as a potential for bias on recovery outcomes. Prior work has demonstrated that people who experience loss of consciousness or amnesia are more likely to seek healthcare(82), but interestingly, the present study found that these factors were *not* associated with poorer recovery. Furthermore, a recent study by Fordal et al(73) found that people who sustained a relatively minor head injury not meeting the criteria for mTBI had comparable symptom presentation and ongoing functional limitations to those who were diagnosed with mTBI when followed up six months after injury. Taken together, it seems that the perceived severity of head injury, based upon commonly accepted diagnostic criteria, does not necessarily influence recovery outcomes. As such, we consider that it is unlikely that this aspect of potential selection bias would affect the findings of this study.

People who had more significant symptoms, or who were perceived to be more likely to participate in the study long-term may have been referred to the study whilst other potential candidates were not, potentially contributing to recruitment bias. As the population characteristics of those who were not referred are unknown, it is not possible to determine whether our sample is a true reflection of all mTBI presentations. However, the demographic characteristics of participants who were referred to the study but not enrolled were comparable to those were enrolled, indicating a lower likelihood of selection bias of our sample.

#### Attrition bias

The attrition rate in this study at three months (29%) and twelve months (41%) is comparable to other longitudinal studies(83), but is a potential source of bias. It is plausible that participants are less likely to be engaged in a study if they are no longer symptomatic(8,53), or conversely, may be lost to follow up if they are particularly symptomatic. In this study, comparative analyses revealed no significant differences in demographic or injury-related characteristics between those who completed all follow-ups and those who did not, except for a lower prevalence of pre-existing mental health issues among individuals who completed all four follow-ups. Pre-existing mental health issues were included as a covariate in multivariable modelling to account for the potential influence of this factor. Thus it is unlikely that the absence of participants lost to follow-up in analysis would have introduced attrition bias(45), although the underrepresentation of individuals with mental health histories could influence symptom burden estimates.

## Conclusion

The findings from this sample of adults with mTBI from a variety of causes provide evidence of the long-term burden of persistent post-concussion symptoms. Of those who completed follow-up, nearly half reported PPCS at twelve months post-injury. Whilst most people returned to work within three months of their injury, a large proportion were found to be working whilst experiencing PPCS. Further exploration of the impact of PPCS on an individual’s employment capacity and capability is essential to support successful and sustained RTW, and to reduce both personal and societal financial implications of mTBI. Baseline assessment of contextual factors and post-concussion symptom severity, particularly symptoms of ‘feeling slow’ and ‘nervousness’, may help clinicians to identify individuals at risk of developing PPCS following mTBI. Early identification of those at greater risk can help develop strategies to improve their outcomes.

## Data Availability

The datasets generated and/or analysed during the current study are not publicly available due to ongoing data analysis as part of the CREST Concussion Recovery Study but are available from the corresponding author on reasonable request.

## Declarations

### Ethics approval and consent to participate

*CREST* was conducted in accordance with the protocol approved by the Royal Perth Hospital Human Research Ethics Committee (#RGS0000003024), Curtin University (HRE2019-0209), University of Western Australia (32019/RA/4/1/7671), Ramsay Health Care (#2009) and St John of God Health Care (#1628), therefore meeting the ethical standards laid down in the 1964 Declaration of Helsinki and its later amendments. All participants provided verbal informed consent prior to participation.

### Competing interests

Prof. Melinda Fitzgerald declares her role as CEO of Connectivity Traumatic Brain Injury Australia Ltd. All other authors of this manuscript have no conflicts of interest associated with this publication.

### Funding

*CREST* was funded through a Neurotrauma Research Program (NRP) grant. SCH is supported by Perron Institute for Neurological and Translation Science partial salary.

### Availability of data and materials

The datasets generated and/or analysed during the current study are not publicly available due to ongoing data analysis as part of the *CREST Concussion Recovery Study* but are available from the corresponding author on reasonable request.

## Supplementary Material

**Supplementary Table 1:** Additional variables collected and examined but not included in ongoing analysis.

| **Participant Characteristics** | **Total**<br>**n=232** |
| --- | --- |
| **Peri-Injury Characteristics** |  |
| **Other Injuries** |  |
| No | 126 (54.5%) |
| Yes | 105 (45.5%) |
| **Legal compensation or litigation** |  |
| No | 201 (88.5%) |
| Yes | 26 (11.5%) |
| **Post-Injury Characteristics** |  |
| **Hospital ED attended** |  |
| RPH | 94 (42.7%) |
| FSH | 17 (7.7%) |
| SCGH | 16 (7.3%) |
| Joondalup | 14 (6.4%) |
| Rockingham | 22 (10.0%) |
| SJOG Midland | 14 (6.4%) |
| SJOG Murdoch | 18 (8.2%) |
| Albany | 17 (7.7%) |
| Geraldton | 7 (3.2%) |
| Other | 1 (0.5%) |
| **CT scan conducted** |  |
| No | 100 (43.9%) |
| Yes | 128 (56.1%) |
| **Initial management** |  |
| ED | 150 (65.8%) |
| Ambulance to ED | 43 (18.9%) |
| ED and GP | 26 (11.4%) |
| GP only | 9 (3.9%) |

**Supplementary Table 2:** Characteristics of eligible participants who were referred to the study but not subsequently enrolled.

| Participant Characteristics | Total |
| --- | --- |
|  | n=136 |
| Age (years), median (IQR) | 29 (22-40) |
| Sex (female) n(%) |  |
| Male | 83 (61.0) |
| Female | 53 (39.0) |
| Hospital ED attended |  |
| RPH | 56 (41.2) |
| FSH | 7 (5.1) |
| SCGH | 13 (9.6) |
| Joondalup | 4 (2.9) |
| Rockingham | 26 (19.1) |
| SJOG Midland | 16 (11.7) |
| SJOG Murdoch | 2 (1.5) |
| Albany | 9 (6.6) |
| Geraldton | 2 (1.5) |
| Other (Self-referred) | 1 (0.7) |

**Supplementary Table 3:** Comparison of those participants who completed all follow-up timepoints (1, 3, 6 and 12 months) to those who had at least one follow-up missing.

|  | Total | Completed all Follow-Ups (n=94) | Follow-ups missing (n=138) | p-value |
| --- | --- | --- | --- | --- |
| **Demographic Factors** |  |  |  |  |
| Age median (IQR) | 33 (24-50) | 36 (25-53) | 31.5 (24-48) | 0.109 |
| Sex n (%) |  |  |  | 0.184 |
| Female | 101 (43.5) | 36 (38.3) | 65 (47.1) |  |
| Male | 131 (56.5) | 58 (61.7) | 73 (52.9) |  |
| **Pre-Injury Factors** |  |  |  |  |
| Previous mTBI n (%) |  |  |  | 0.282 |
| Yes | 118 (51.1) | 44 (46.8) | 74 (54.0) |  |
| No | 113 (48.9) | 50 (53.2) | 63 (46.0) |  |
| Previous mental health issues n (%) |  |  |  | **0.030\*** |
| Yes | 83 (35.8) | 25 (26.6) | 58 (42.0) |  |
| No | 148 (63.8) | 68 (72.3) | 80 (58.0) |  |
| **Peri-Injury Factors** |  |  |  |  |
| Mechanism of injury n (%) |  |  |  | 0.249 |
| Sport Related | 74 (31.9) | 34 (36.2) | 40 (28.9) |  |
| Non-sport related | 158 (68.10) | 60 (63.8) | 98 (71.1) |  |
| Loss of Consciousness |  |  |  | 0.939 |
| Yes | 66 (28.4) | 27 (28.7) | 39 (28.3) |  |
| No | 124 (53.5) | 51 (54.3) | 73 (52.9) |  |
| Unsure | 42 (18.1) | 16 (17.0) | 26 (18.8) |  |
| **Post-Injury Factors** |  |  |  |  |
| PCSS Symptom Severity median (IQR) | 29 (16-49) | 25 (14-49) | 30.5 (17-49) | 0.099 |

**Supplementary Figure 1.**
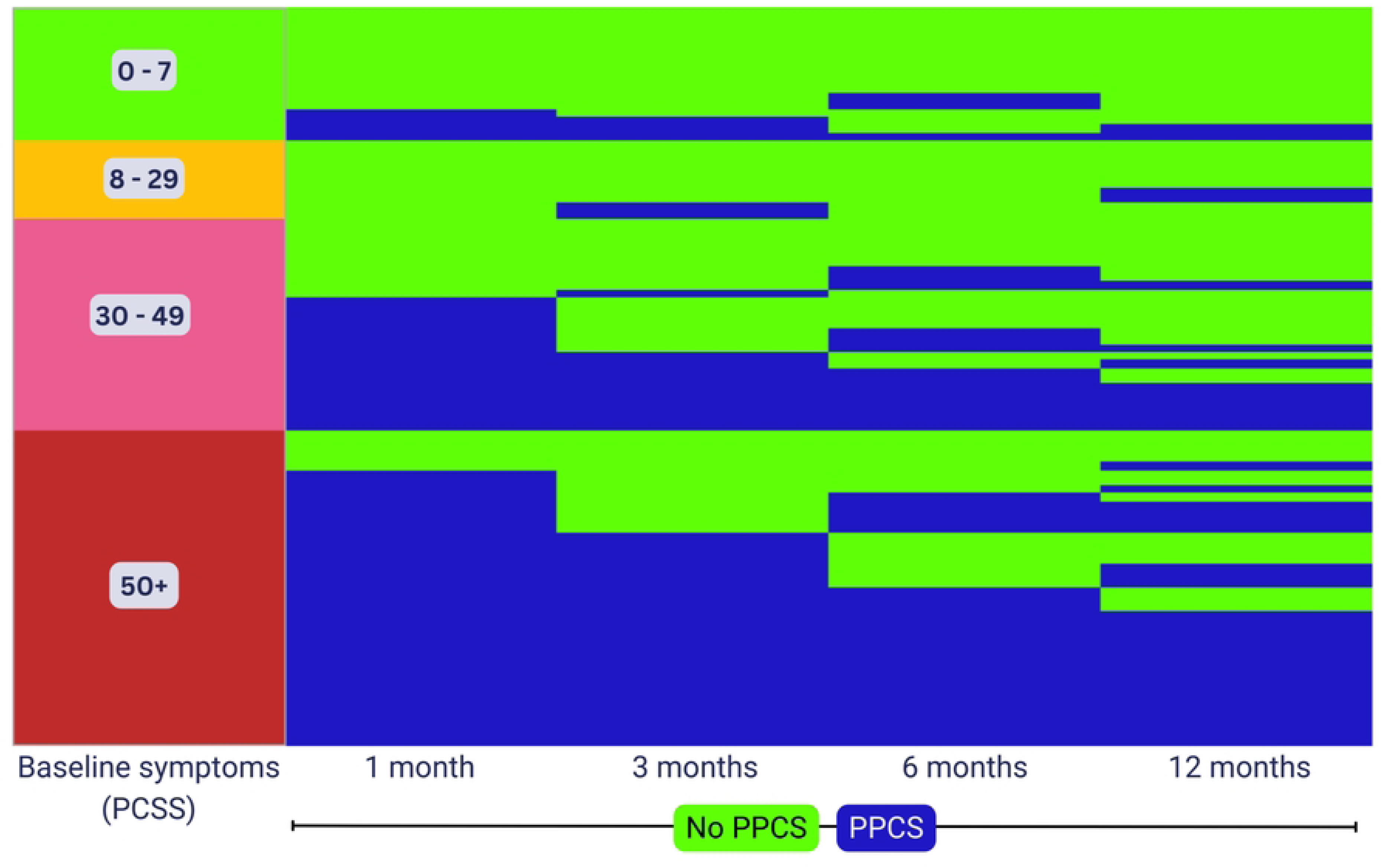
Relationship between PCSS symptom severity score at initial enrolment and presence or absence of PPCS at each timepoint. Each row represents one participant who completed all follow-up timepoints (n=94).

## Notes

### Clinical Trial

The Concussion REcovery STudy (CREST) was registered with the Australian New Zealand Clinical Trials Registry (ACTRN12619001226190).

### Clinical Protocols

https://bmjopen.bmj.com/lookup/doi/10.1136/bmjopen-2020-046460

### Author Declarations

CREST was conducted in accordance with the protocol approved by the Royal Perth Hospital Human Research Ethics Committee (#RGS0000003024), Curtin University (HRE2019-0209), University of Western Australia (32019/RA/4/1/7671), Ramsay Health Care (#2009) and St John of God Health Care (#1628), therefore meeting the ethical standards laid down in the 1964 Declaration of Helsinki and its later amendments. All participants provided verbal informed consent prior to participation.

